# The Impact of Policy Restrictions and Mobility Changes on Excess Mortality During the COVID-19 Pandemic in The Netherlands, 2020-2022

**DOI:** 10.1101/2025.03.25.25324481

**Authors:** Dimiter Toshkov, Camila Caram-Deelder, Brendan Carroll, Frits R. Rosendaal

## Abstract

We analyzed the impact of the COVID-19 policy restrictions on mobility patterns and excess mortality at the regional level in The Netherlands between 2020 and 2022. Our analysis combines data on public policies, mobility patterns from the Google Mobility Reports, officially registered COVID-19 cases and deaths, and region-specific measures of excess mortality over a relatively long time period extending beyond the first wave of the pandemic. We modeled the dynamic relationships of these variables as a system in which policy responds to information about the pandemic; mobility reacts both to information about the pandemic and to policy; the number of COVID-19 cases is influenced by changes in mobility and policy; and excess mortality is affected directly by the policy restrictions and indirectly via the impact of policy on mobility. The results confirm that the stringency of policy restrictions increased with the number and growth rates of COVID-19 cases and deaths. Mobility, as reflected in presence in public places (transport hubs, retail, work), decreased while presence at residential locations increased in response to stricter policies and higher COVID-19 case and death counts in preceding weeks. The number of new COVID-19 cases declined when stricter policy restrictions were enacted and reduced presence in public places (following a two-week lag). Excess mortality decreased with stricter policy restrictions (with a five-week lag) and, to a lesser extent, with reduced presence in public places and increased presence in places of residence. Importantly, the effects of policy restrictions and mobility diminished with consecutive COVID-19 waves. Overall, the evidence shows that policy restrictions were effective in limiting the spread of the pandemic and in saving lives. While policies influenced mobility patterns, their impact was not fully mediated by mobility changes.

## Introduction

Starting in early 2020, governments around the world enacted restrictive policies (non-pharmaceutical interventions) with the aim to curb and slow down the spread of the COVID-19 pandemic. These policies included physical distancing, face mask requirements, school closures, restrictions on large gatherings, recommendations to work from home, bans on use of public spaces such as parks, reduced opening hours of shopping centres, and – ultimately – lockdowns and curfews (1). These policies imposed significant costs on societies, including limits on personal liberties and financial losses for businesses. At the same time, the restrictions may have saved lives by containing the spread of the coronavirus, protecting the capacity of hospitals, and limiting the reach of COVID-19. It is therefore imperative to assess their impact on excess mortality, as one crucial indicator of the toll of the pandemic.

There are a significant number of studies that have attempted to assess the impact of the policy restrictions on health outcomes (i.a. 2–12). On balance, existing research finds strong evidence that restrictive measures reduced the number of infections and the number of deaths related to COVID-19, although there is heterogeneity in the effects of different policies and context-specificity of these effects (for systematic reviews, see 13–15).

While many studies address the question of COVID-19 policy impact, the vast majority of these studies focus on the first wave of the pandemic and the government responses to it. Yet, the effectiveness of measures may have varied over the different waves of the pandemic. The effectiveness may have varied due to different levels of compliance (e.g., ‘pandemic fatigue’) and – as a result – different effects on the mediator ‘mobility’. Policy effects might vary because of changes in the mix of policy measures over time as well. Furthermore, even studies that evaluated the impact on the death toll of the pandemic often focused on the number of COVID-19 related deaths as reported by the governments, while excess mortality is a more appropriate measure (16–21). This is because the number of deaths registered as caused by COVID-19 does not accurately represent the actual death toll of the pandemic. and because of regional and temporal variation in the decisions leading to classifying a death as COVID-19 related or not. There are also a limited number of studies that explore within-country variation in excess mortality, while having regional-level data provides opportunities to assess the indirect policy impact mediated by changes in mobility.

In this article, we assess the impact of the policy measures by examining the dynamic relationships between policy, changes in patterns of mobility over time and across regions within a country, changes in the dynamic of the pandemic (which may affect the level of policy restrictions and mobility directly) and, ultimately, excess mortality. One of our assumptions is that changes in mobility (people spending more time at home and less time in public spaces such as transport hubs, shopping centres and offices) provide the main mechanism through which the public policies could have affected COVID-19-induced excess mortality. Hence, if we are able to identify the effect of public policies on mobility and of mobility on excess mortality, we can provide an assessment of the effect of the policies on mortality as well. In sum, our main objective is to provide an assessment of the direct and indirect impact of restrictive policy measures on excess mortality. The main research questions are (1) what was the effect of policy restrictions on mobility, (2) what was the effect of changes in mobility on excess mortality, (3) what was the effect of restrictive policy measures on excess mortality overall and specifically via reducing mobility?

We study these questions focusing on The Netherlands in the period from 2020 to 2022, with variation over time and between the 12 regional units (provinces) in the country. During this period, various policy restrictions were imposed, modified and eventually lifted. While there was limited variation in the presence or absence of restrictive policies across provinces in the country, compliance with the policies, as reflected in changes in mobility patterns, may have differed. This variation allows us to estimate the indirect effect of the policy restrictions, as recommended in recent work (22).

The causal effect of the policies is difficult to assess because the imposition of restrictions followed the course and severity of the pandemic: restrictive policies were more likely to be adopted when COVID-19 cases surged, which in itself leads to increases in mortality. Furthermore, there is not much variation in the timing and nature of restrictions imposed *within* most individual countries (and only to some extent across countries). These associations and lack of variation, as well as related concerns about confounding, are major challenges for the identification of the causal impact of the COVID-19 policy measures.

Various research designs and methodologies have been employed to assess the effects of the policies. For example, Méndez-Lizárraga et al. (4) use change-point analysis. Chernozhukov et al. apply econometric techniques to panel data from US states (2) and US counties (3). Askitas et al. (5) use daily data from 175 countries to estimate the dynamic effects of different policy interventions.. Some studies focus only on the relationship between mobility changes and health outcomes (assessed with correlation coefficients) (e.g. 23), others use changes in mobility as a proxy for policy (e.g. 4), while the most sophisticated incorporate mobility in the models designed to assess the policy impact (2,3).

The main strengths of our approach compared to the existing literature are that (a) we use a highly disaggregated (weekly province-level) estimate of excess mortality, which provides a reliable and fine-grained measure of the death toll of the pandemic(16–20,24); (b) we combine policy, mobility and COVID-19-related data so that we can assess simultaneously various direct and indirect paths of influence; and (c) we analyse an extended period of time that goes beyond the first wave of the pandemic, which is the focus of the vast majority of existing studies. This is important, as the effects of policy and mobility might have changed considerably over the different waves of the pandemic. In addition, in our statistical models we control for important potential confounders of these relationships, including demographic variables related to the population structure of the provinces, as well as data on the weather that might have affected both mobility patterns and mortality.

Our study found that policy stringency increased in response to higher numbers and growth rates of COVID-19 cases and deaths, with the impact of deaths increasing over the course of the pandemic. Stricter policy in turn reduced citizens’ mobility, while deaths (and to a lesser degree cases) also had a direct negative effect on mobility. Policy restrictions and changes in mobility had significant effects on the number of cases after a lag of two weeks, while only the former significantly affected excess mortality in a consistent way. Importantly, our study showed that while the effects of policy restrictions and mobility weakened with successive waves of the pandemic, overall, the restrictions were effective in reducing mobility and saving lives.

### Theoretical causal mechanisms

In this section we outline the theoretical model that we use to guide the empirical analysis. The model incorporates the following hypothesized relationships. First, as the pandemic unfolds, information about the state of the spread and impact of COVID-19 affects the likelihood that restrictive policy measures area adopted (and later on lifted). The most policy-relevant information signals have been the number of new COVID-19 cases and deaths, as well as their growth rates. Policy stringency is expected to increase with a higher number and higher growth rates of COVID-19 cases and deaths, and to decrease when fewer cases and deaths are registered and the pandemic subdues.

Pandemic-related information might also impact mobility patterns directly, i.e., not as a result of restrictions, as citizens will tend to spend more time at home and avoid public places when higher numbers of COVID-19 cases and deaths are reported for fear of the virus. Mobility might also respond to the course of the pandemic via the path Information > Policy restrictions > Mobility changes. Many policy measures limited presence in public places by imposing physical distancing, school closures, bans on large gatherings, work-from-home recommendations, restricting access to parks and shopping centres and total societal lockdowns. We hypothesize that restrictive policy measures affect mobility patterns immediately when imposed without a lag (at the week level of aggregation).

Next, policy restrictions and changes in mobility affect the number of COVID-19 cases, but with a lag of *j* weeks (for example, two weeks). Some of the policy effects are expected to be exercised via changes in mobility. But it is also possible that restrictive policy measures affect the spread of the pandemic via alternative mechanisms, for instance improved ventilation in buildings and face mask requirements, or testing. Finally, changes in the number of COVID-19 cases lead to COVID-19 related deaths and excess mortality, with a lag of *k* weeks (for example, four weeks).

As COVID-19 cases and growth affect policy (and mobility) and, in their turn, policy and mobility affect the number of new COVID-19, it would seem that our model contains a cyclic causal relationship, which presents a challenge for causal identification. But the temporal ordering and lag structure that we impose (mobility and policy react instantaneously to changes in the COVID-19 growth rate, but affect cases only after a lag of time) provides a solution to this challenge (cf. 2,3). For a different approach, see (25) who attempt to learn the causal structure of this system of variables inductively via machine learning.

## Materials and methods

The study is based on an observational time-series cross-sectional design that exploits variation in the imposition of public policy measures over time and variation in the changes in mobility patterns over time *and* across spatial units. Hence, the unit of observation is a province in a week. The spatial unit is the province level in The Netherlands (N_provinces_=12). The provinces differ significantly when it comes to population density, from in 187 people per km^2^ in Drenthe to 1374 in South Holland (see the Supplementary material for details). The time scope of the analysis is between third week of February 2020 and the second week of October 2022, which is the last date of availability for the mobility data (N_weeks_=140).

We translate our theoretical model in the following system of equations:

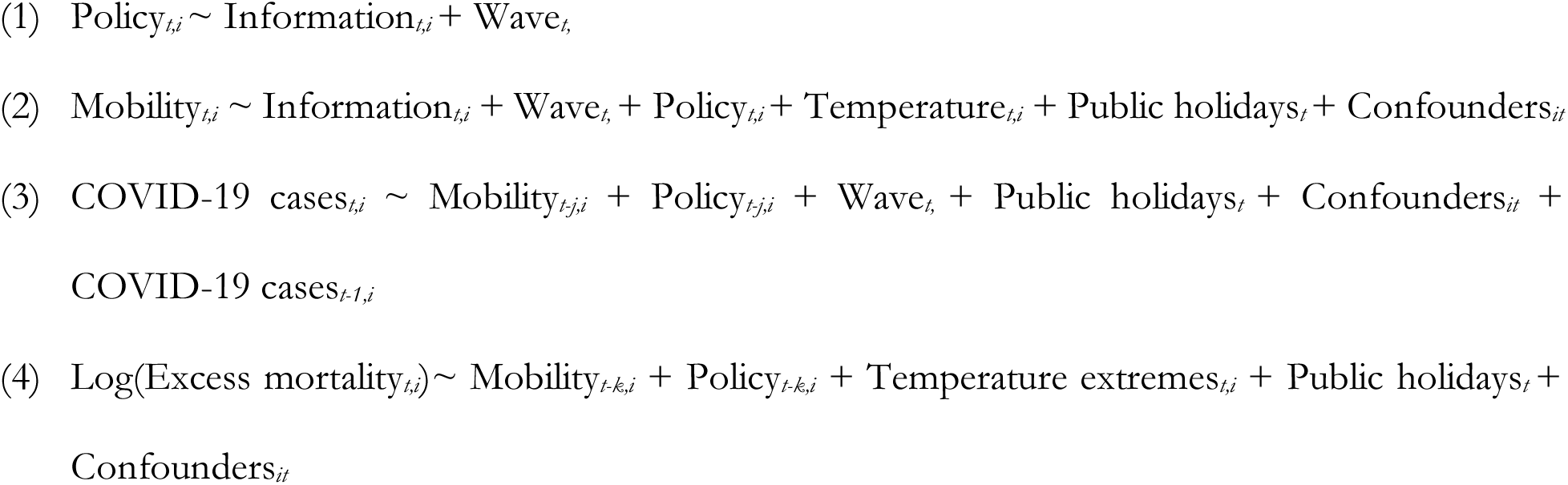

In the equations above, *t* is a week indicator and *i* is a spatial unit indicator. *j* is the indicator lag time for the measurement of number of cases and *k* is the lag time for the policy. The effect of the public policy restrictions via mobility is given by the product of the coefficient of Policy in equation 2 and the coefficient of changes in Mobility in equation 3, for COVID-19 cases and in equation 4 for excess mortality.

We use linear regression to estimate the equations. Where appropriate, we add fixed effects for the provinces, and we cluster the standard errors in estimating equation 1 at the province-level. In addition, we explore interactions of the main variables of interest with the COVID-19 pandemic wave and across the spatial units.

The variable ‘Policy’ is operationalized as the general policy stringency index constructed from the Oxford COVID-19 Government Response Tracker (26). The variable is aggregated per week by using the mean for the week. We rescale the variable from the original 0-to-100 range to the 0-to-10 range. The variable tracking pandemic-related ‘Information’ is operationalized as the natural logarithm (the log) of the number of new province-specific weekly COVID-19 cases and the number of new province-specific weekly COVID-19 related deaths. To make the effects of these two indicators comparable, we scale the number of cases by dividing it by 1,000 (prior to taking the log). We use the seven-day moving average of the daily values of cases and deaths and then aggregate per week by taking the mean for the seven days of the week.

The variable ‘Mobility’ is operationalized as an index constructed from the Google Mobility Report, which tracks for each date the percentage change in presence at six different categories of places in a province relative to a baseline period in January-February 2020 in the same province. We take the seven-day moving average of the daily values and then aggregate per week by taking the mean for the seven days of the week. We focus on mobility measures about four particular kinds of places (i.e. places of work, transport hubs, grocery shops and markets, and residential places), as reported by the Google Mobility Report.

‘Excess mortality’ is operationalized as the number of deaths in a province in a week relative to the baseline in 2019 in the province and the week. We use Poisson regression analysis to estimate excess mortality in terms of incidence rate ratios (IRR) for each week 2020, 2021 and 2022 compared with the baseline year (2019). The models are based on individual-level register data for the entire Dutch population provided via a special arrangement by the CBS (Statistics Netherlands). The measure adjusts for the composition of the population in the province in terms of sex, age, household income and immigration background, and for the calendar month as well. We take the log of the IRR. More details about the estimation and the mathematical models behind the estimates is provided in (21). Figure S1 in the Supporting Information shows the descriptive trends in these main variables of interest.

Our models also include some additional covariates that capture additional influences on mobility, COVID-19 cases and excess mortality. Public holidays is a variable that is necessary to include because of the way mobility changes are measured (relative to a baseline in February). We would expect that this variable picks up the effect of reductions in mobility that are not related to COVID-19 or policy responses, but to other relevant events. The variable indexes the weeks of Christmas, New Year and Easter. The variables related to Temperature track the average weekly temperature (measured at a weather station within each province) for the equations of Mobility and COVID-19 cases and whether the weekly average of the minimum temperature was below zero or above 25 degrees Celsius in the equation of excess mortality. The demographic covariates relate to the composition of the population of the province in a week. In particular, we track the share of old people (65+), the share of women, the share of low and low-middle income households and the share of 1^st^ generation immigrants. The variable has some variation within provinces over time, but the extent of this variation is limited.

All models reported below are estimated using the R software environment for statistical computing and graphics using base R and the *fixest* package (27) for the models reported in Table 1. The figures of marginal effects are produced with the packages *modelsummary* (28) and *coefplot* (*29*).

**Table 1.** Policy stringency as a function of COVID-19 cases and deaths, per COVID-19 wave.

|  | Level of<br>Policy stringency index |  | Change in<br>Policy stringency index |  |
| --- | --- | --- | --- | --- |
|  | Model 1a | Model 1b | Model 1c | Model 1d |
| (Intercept) | 3.43 [2.99, 3.87]<br>p<0.01 *** | 3.16 [2.34, 3.98]<br>p<0.01 *** | 0.13 [0.11, 0.15]<br>p<0.01 *** | 0.09 [0.05, 0.12]<br>p<0.01 *** |
| COVID-19 cases<br>- log (number)<br>- growth rate | 0.35 [0.26, 0.43]<br>p<0.01 *** | 0.45 [0.25, 0.64]<br>p<0.01 *** | 0.31 [0.25, 0.36]<br>p<0.01 *** | 0.57 [0.43, 0.70]<br>p<0.01 *** |
| COVID-19 deaths<br>- number (lag 1)<br>- growth rate | 0.01 [0.00, 0.02]<br>p<0.01 ** | 0.01 [0.00, 0.01]<br>p=0.02 * | 0.01 [0.00, 0.01]<br>p<0.01 *** | 0.01 [0.01, 0.01]<br>p<0.01 *** |
| Second wave<br>(indicator) | -0.54 [-0.88, -0.21]<br>p<0.01 ** | -0.03 [-0.96, 0.90]<br>p=0.95 | -0.18 [-0.20, -0.17] p<0.01 *** | -0.13 [-0.16, -0.09] p<0.01 *** |
| Third wave<br>(indicator) | -4.32 [-4.84, -3.80]<br>p<0.01 *** | -3.42 [-4.19, -2.65]<br>p<0.01 *** | -0.16 [-0.18, -0.15] p<0.01 *** | -0.12 [-0.16, -0.09] p<0.01 *** |
| COVID-19 cases<br>× Second wave |  | -0.13 [-0.30, 0.04]<br>p=0.12 |  | -0.43 [-0.58, -0.27] p<0.01 *** |
| COVID-19 cases<br>× Third wave |  | -0.21 [-0.37, -0.05]<br>p=0.01 * |  | -0.52 [-0.66, -0.37] p<0.01 *** |
| COVID-19 deaths<br>× Second wave |  | 0.00 [0.00, 0.01]<br>p=0.33 |  | 0.00 [-0.01, 0.00]<br>p<0.01 *** |
| COVID-19 deaths<br>× Third wave |  | 0.05 [0.03, 0.06]<br>p<0.01 *** |  | 0.00 [-0.01, 0.00]<br>p=0.05 + |
| *Num.Obs.* | *1644* | *1644* | *1644* | *1644* |
| *R2 Adj.* | *0.624* | *0.639* | *0.121* | *0.172* |

*The numbers show the unstandardized coefficients from linear regression models, which indicate the implied change in the Policy stringency index for a one-unit change in the covariate. Standard errors are clustered at the level of provinces (N=12). 95% Confidence intervals are reported in the square brackets. Significance levels of p values: \*\*\* < 0.001; \*\* < 0.01; \* < 0.05; + < 0.10. Exact p values when > 0.01 are printed.*

## Results

### Part I: Policy restrictions as a function of COVID-19 cases and reported deaths

In the first part of the empirical analysis, we look at the stringency of the policy restrictions in force as a function of the pandemic-related information. Models 1a and 1b in Table 1 analyse *the level* of policy restrictions in place as a function of the (logged) number of the newly-registered COVID-19 cases and the number of deaths (in the preceding week). Models 1c and 1d analyse *the change* in the policy stringency index as a function of the growth rate in COVID-19 cases^5^ and deaths. As the data are aggregated at the weekly level, we expect that the number of cases in the same week and the number of deaths in the preceding week are most relevant for the level of policy restrictions in force.

Table 1 shows the results of two linear regression models for each of the two outcomes of interest (levels and changes in the policy): one *with* and one *without* interactions between the predictors and the three waves of the pandemic in the period 2020-2022. In all four models the standard errors are clustered at the level of the province, because there is no variation in the policy data at that level.

As expected, both the number of cases and the number of deaths are significant predictors of the stringency of the public policy in place in the subsequent week (Models 1a and 1b). The effect of the number of cases was strongest (largest coefficient) during the first wave, but declined significantly in the subsequent two waves. At the same time, the effect of the number of deaths remained constant during the first two waves and even increased during the third wave. Figure 1 provides an illustration.

**Fig 1.**
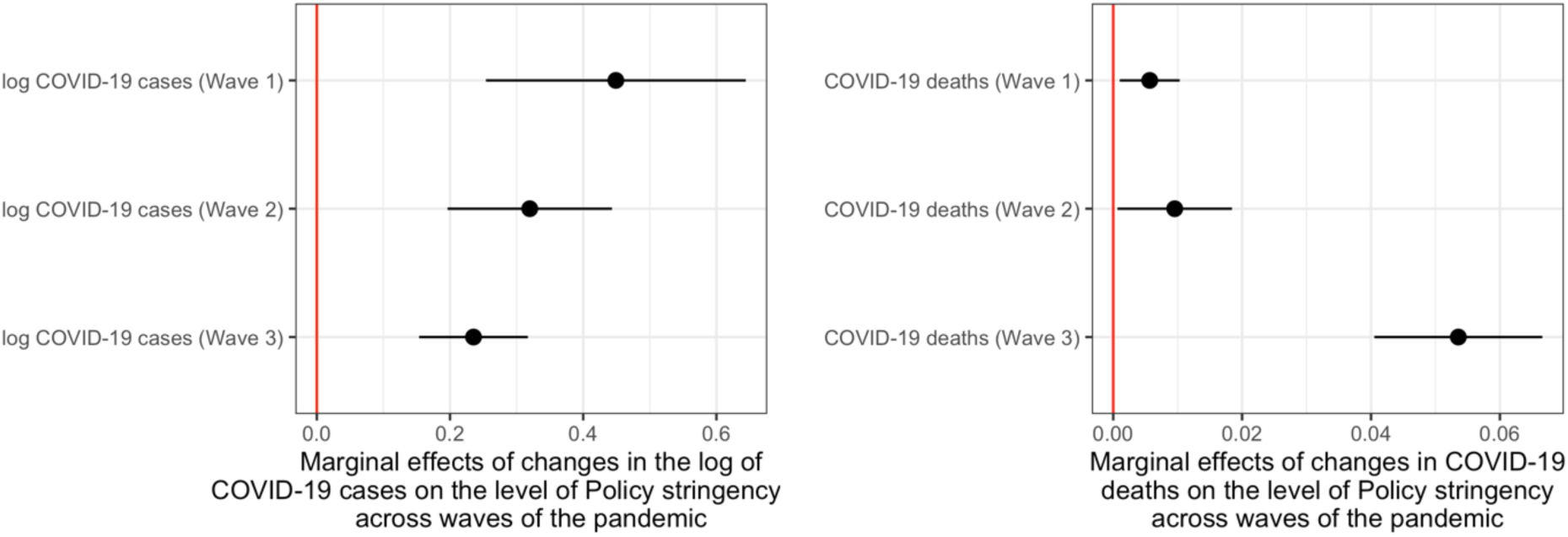
Marginal effects (point estimates and 95% confidence intervals) of the number of COVID-19 cases and deaths on the level of Policy stringency in The Netherlands, across the first three waves of the pandemic (2020-2022) The figure shows the predicted effects of one-point increases in the log of COVID-19 cases (left panel) and COVID-19 deaths (right panel) on the level of Policy stringency during each of the three stages of the pandemic in the period 2020-2022 in The Netherlands. One-point increases in the log of the number of cases and deaths correspond approximately to doubling the number of cases and deaths. Results based on Model 1b in Table 1.

The growth rate of cases and deaths also predict changes in the policy stringency index. The same pattern of declining effect for cases but increasing effect of deaths during the third wave can be observed in Models 1c and 1d.

### Part II: Mobility as a function of COVID-19 cases and policy restrictions

Table 2 presents four linear regression models that analyse changes in mobility. Each of the four models focuses on a different aspect of mobility. The dependent variable is the percentage change in the presence of people in particular types of places compared with a baseline period in the pre-pandemic period in early 2020. All four models include fixed effects at the level of the province, as well as control variables related to the demographic structure of the population in the province (which has some minor variation within-provinces over time as well).

**Table 2.** Changes in mobility as a function of policy stringency, COVID-19 cases and deaths, and additional covariates.

|  | **Model 2a**<br>**Places of**<br>**work** | **Model 2b**<br>**Public**<br>**transport hubs** | **Model 2c**<br>**Grocery shops**<br>**and markets** | **Model 2d**<br>**Residential**<br>**places** |
| --- | --- | --- | --- | --- |
| Policy stringency | -2.74 [-3.03, -2.46] p<0.01 *** | -3.14 [-3.50, -2.79] p<0.01 *** | -0.51 [-0.80, -0.22] p<0.01 *** | 0.96 [0.89, 1.02] p<0.01 *** |
| COVID-19 cases (log, lag 1) | 0.05 [-0.33, 0.42] p=0.81 | -2.30 [-2.77, -1.83] p<0.01 *** | -0.96 [-1.34, -0.58] p<0.01 *** | 0.41 [0.32, 0.49] p<0.01 *** |
| COVID-19 deaths (log, lag 1) | -1.39 [-1.86, -0.93] p<0.01 *** | -1.44 [-2.02, -0.85] p<0.01 *** | -0.61 [-1.09, -0.13] p=0.01 * | 0.61 [0.51, 0.72] p<0.01 *** |
| Second wave | 4.50 [2.96, 6.03] p<0.01 *** | 11.28 [9.36, 13.20] p<0.01 *** | 4.86 [3.30, 6.42] p<0.01 *** | -3.37 [-3.72, -3.02] p<0.01 *** |
| Third wave | -3.56 [-5.93, -1.20] p<0.01 ** | 14.97 [12.01, 17.92] p<0.01 *** | 10.55 [8.15, 12.95] p<0.01 *** | -3.41 [-3.95, -2.87] p<0.01 *** |
| Public holidays | -10.51 [-12.07, -8.95] p<0.01 *** | -4.87 [-6.82, -2.93] p<0.01 *** | -5.45 [-7.04, -3.86] p<0.01 *** | 1.82 [1.46, 2.17] p<0.01 *** |
| Max. temperature | -0.43 [-0.50, -0.36] p<0.01 *** | 0.34 [0.25, 0.42] p<0.01 *** | 0.38 [0.31, 0.46] p<0.01 *** | -0.08 [-0.10, -0.07] p<0.01 *** |
| *Num.Obs.* | *1626* | *1622* | *1630* | *1632* |
| *R2 Adj.* | *0.519* | *0.798* | *0.664* | *0.859* |

*The numbers show the unstandardized coefficients from linear regression models, which indicate the implied change on Mobility (defined as the percentage change in the presence of people compared with a baseline period in the pre-pandemic period in early 2020 in particular types of places) for a one-unit change in the covariate. The models include fixed effects at the province level (N=12), as well as controls for the demographic structure of the provinces (share of 65+, share of women, share of low-income households, share of 1<sup>st</sup> generation immigrants). 95% Confidence intervals are reported in the square brackets. Significance levels of p values: \*\*\* < 0.001; \*\* < 0.01; \* < 0.05; + < 0.10. Exact p values when > 0.01 are printed.*

The policy stringency index has statistically significant negative associations with presence in places of work, public transport hubs and grocery shops and markets; it has a positive association with presence in residential places. This implies that mobility responded to public policy, and people reduced their presence in work, transport and shopping places and stayed more at home, in accordance with the policy restrictions. The effect of public policy comes on top of any direct responses of the population to the state of the pandemic, as captured by the information variables related to the number of cases and deaths.

The number of deaths has significant negative associations with mobility, so that in the aftermath of weeks with more COVID-19-related deaths people spent less time in places of work, public transport hubs and grocery shops and markets, and more time in residential places. The number of reported cases also has the expected effects, except on Places of work, where the effect is not significant.

We also note that the variables for the weeks with big public holidays (Christmas and Easter) and the maximum average temperature of the week in the province have the expected associations with changes in mobility, with people spending more time at home and less time in other places during holidays and less time at home and at the office when it is warmer.

When we include interaction effects between the policy stringency index and the COVID-19 waves, we can see that the effects of policy on mobility are substantially attenuated during the second and the third wave, but remain significant and in the direction reported in Table 2. The results of these models are reported in the Supporting Information (Table S1). These models also show that the number of COVID-19 cases has the expected effect only during the first wave, but not after (which explains the overall lack of a consistent effect of this variable in Table 2). The number of deaths has the strongest effect in the expected direction during the second wave of the pandemic.

We also explored possible differential effects of policy and COVID-19-related information on mobility across different provinces. The main results of these models with respect to the effects of policy stringency are presented in Figure 2. The figure shows that the negative effect of policy stringency on mobility related to places of work was greatest in Limburg and Gelderland; with regard to transport hubs, it was greatest in Noord-Holland; with regard to grocery shop and markets, it was variable but greatest in Limburg and Zeeland; and with regard to places of residence, the positive effect of policy stringency was greatest in Zeeland, but the variation across provinces was minor.

**Fig 2.**
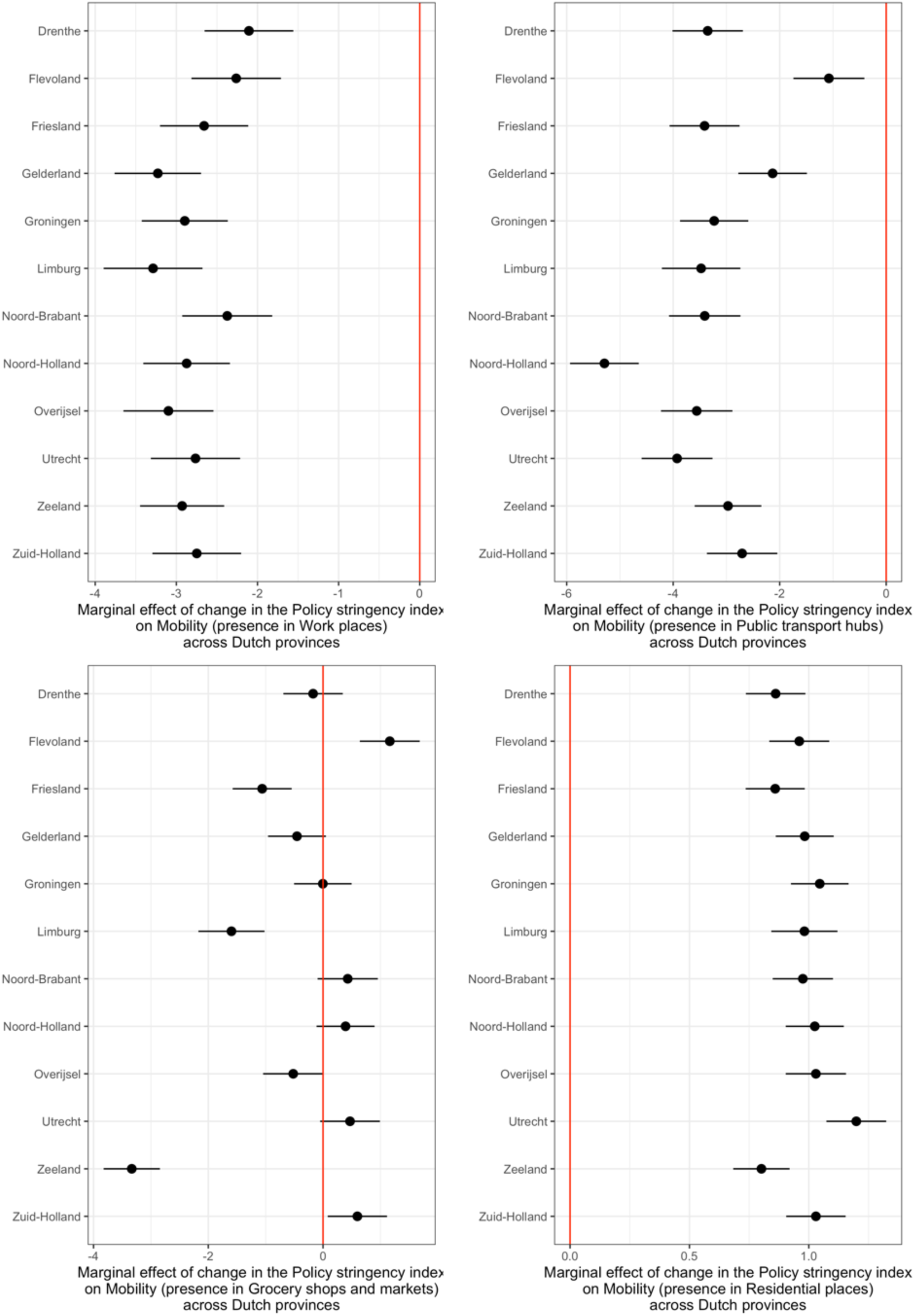
Marginal effects of changes in the Policy stringency index on changes in mobility for four types of places across provinces in The Netherlands, 2020-2022 The figure shows the predicted marginal effects ((point estimates and 95% confidence intervals) of one-point increases in the Policy stringency index on Mobility (defined as the percentage change in the presence of people compared with a baseline period in the pre-pandemic period in early 2020) in four types of places (Work places, Public transport hubs, Grocery shops and markets, and Residential places) depicted in the four panels of the figures, in each of the 12 provinces in The Netherlands. Results based on variations of the models reported in Table 2 with added interactions of policy stringency and province indicators.

### Part III: COVID-19 cases as a function of policy restrictions and mobility changes

Next, we look at the number of COVID-19 cases as a function of the lagged value of the number of COVID-19 cases, the lagged value of policy stringency, and the lagged values of the changes in mobility. All linear regression models reported in Table 3 also include province fixed effects and demographic controls for the population structure, in addition to fixed effects for the pandemic wave, public holidays and a variable tracking the average maximum temperature in the province.

**Table 3.** Number of registered COVID-19 cases (logged) as a function of policy stringency, changes in mobility and additional covariates.

|  | **Model 3a**<br>**COVID-19**<br>**cases (log)** | **Model 3b**<br>**COVID-19**<br>**cases (log)** | **Model 3c**<br>**COVID-19**<br>**cases (log)** | **Model 3d**<br>**COVID-19**<br>**cases (log)** | **Model 3e**<br>**COVID-19**<br>**cases (log)** |
| --- | --- | --- | --- | --- | --- |
| Lagged<br>COVID-19<br>cases (log) | 0.83 [0.81, 0.86]<br>p<0.01 *** | 0.83 [0.81, 0.85]<br>p<0.01 *** | 0.85 [0.83, 0.87]<br>p<0.01 *** | 0.84 [0.82, 0.86]<br>p<0.01 *** | 0.85 [0.82, 0.87]<br>p<0.01 *** |
| Policy<br>stringency<br>(lag 1) | -0.08 [-0.09,<br>-0.06] p<0.01 *** | -0.06 [-0.08,<br>-0.04] p<0.01 *** | -0.06 [-0.08,<br>-0.04] p<0.01 *** | -0.08 [-0.09,<br>-0.06] p<0.01 *** | -0.05 [-0.08,<br>-0.03] p<0.01 *** |
| Work (lag 2) |  | 0.01 [0.00, 0.01]<br>p<0.01 *** |  |  |  |
| Transport<br>(lag 2) |  |  | 0.00 [0.00, 0.01]<br>p<0.01 *** |  |  |
| Retail (lag 2) |  |  |  | 0.00 [0.00, 0.01]<br>p=0.05 * |  |
| Residence<br>(lag 2) |  |  |  |  | -0.02 [-0.03,<br>-0.01] p<0.01 ** |
| Public<br>holidays | 0.01 [-0.09, 0.10]<br>p=0.92 | 0.00 [-0.09, 0.10]<br>p=0.96 | 0.01 [-0.08, 0.11]<br>p=0.81 | 0.00 [-0.09, 0.10]<br>p=0.93 | 0.01 [-0.09, 0.10]<br>p=0.90 |
| Max.<br>temperature | -0.04 [-0.04,<br>-0.03] p<0.01 *** | -0.03 [-0.04,<br>-0.03] p<0.01 *** | -0.04 [-0.04,<br>-0.03] p<0.01 *** | -0.04 [-0.04,<br>-0.03] p<0.01 *** | -0.04 [-0.04,<br>-0.03] p<0.01 *** |
| Second wave | 0.44 [0.35, 0.53]<br>p<0.01 *** | 0.42 [0.33, 0.51]<br>p<0.01 *** | 0.38 [0.29, 0.48]<br>p<0.01 *** | 0.42 [0.33, 0.51]<br>p<0.01 *** | 0.36 [0.26, 0.47]<br>p<0.01 *** |
| Third wave | 0.41 [0.27, 0.55]<br>p<0.01 *** | 0.43 [0.29, 0.57]<br>p<0.01 *** | 0.33 [0.19, 0.48]<br>p<0.01 *** | 0.37 [0.22, 0.51]<br>p<0.01 *** | 0.33 [0.18, 0.48]<br>p<0.01 *** |
| *Num.Obs.* | *1632* | *1626* | *1622* | *1630* | *1632* |
| *R2 Adj.* | *0.957* | *0.957* | *0.957* | *0.957* | *0.957* |

*The numbers show the unstandardized coefficients from linear regression models, which indicate the implied change on the log of the number of COVID-19 cases for a one-unit change in the covariate. The models include fixed effects at the province level (N=12), as well as controls for the demographic structure of the provinces (share of 65+, share of women, share of low-income households, share of 1<sup>st</sup> generation immigrants). 95% Confidence intervals are reported in the square brackets. Significance levels of p values: \*\*\* < 0.001; \*\* < 0.01; \* < 0.05; + < 0.10. Exact p values when > 0.01 are printed.*

As expected, the number of COVID-19 cases is strongly auto-correlated, as indicated by the significant lagged value of this variable and the adjusted R-squared for the models. More importantly, the lagged value of the policy stringency index has negative association with the number of COVID-19 cases, implying that the policy restrictions worked to reduce the spread of the pandemic. The variables tracking different type of mobility changes also have significant effects (with one exception). More time at work or transport places (or, equivalently, smaller reductions in presence at such places compared to the baseline) is associated with more COVID-19 cases two weeks after. More presence at home (residential places) has a negative association with the number of COVID-19 cases. We obtain substantively the same results when we model the growth rate in cases rather than the level.

When we include interactions between the effects of policy stringency and mobility with COVID-19 waves, we find evidence that the effects varied over the course the pandemic (see Table S2 in the Supporting Information). Figure 3 illustrates the results with respect to policy. The negative effect of policy restrictions was strongest during the second wave, while it was weakest – and not estimated precisely to be different from zero – during the third wave. Mobility changes related to work and transport places had significant effects mostly during the first wave. There is not much variation in the effect of policy across provinces.

**Fig 3.**
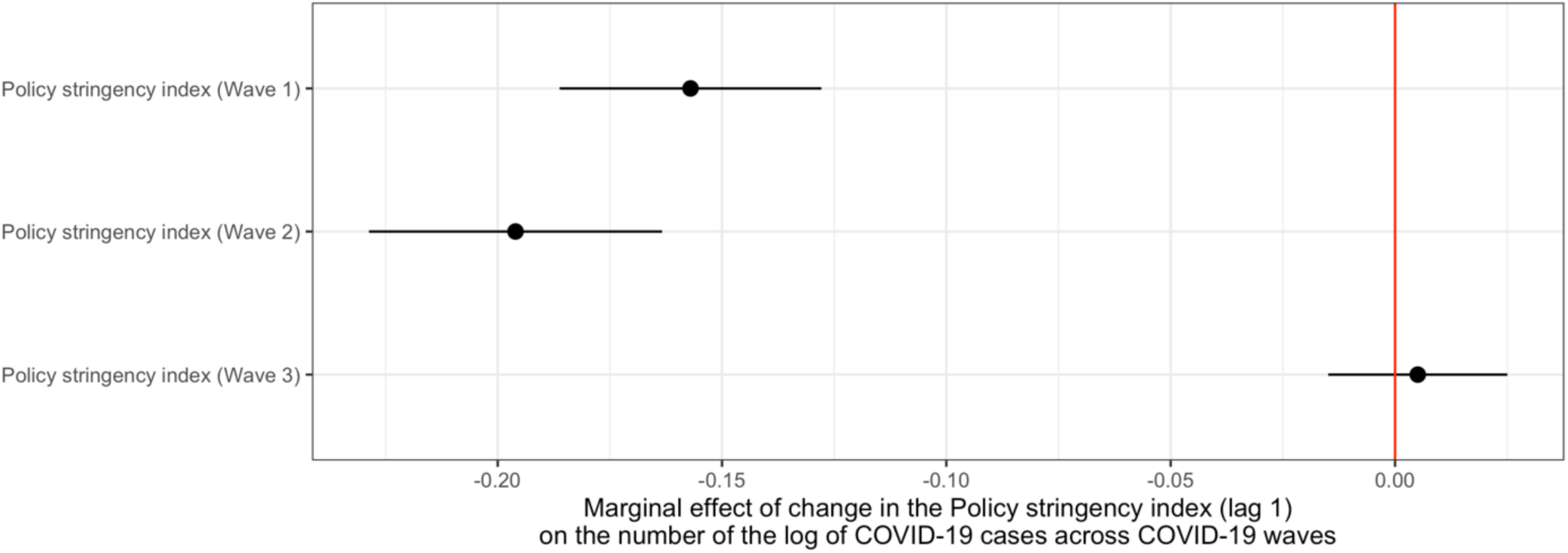
Marginal effects of changes in the Policy stringency index (lag 1) on the log of the number of COVID-19 cases in The Netherlands, across the first three waves of the pandemic (2020-2022) The figure shows the predicted marginal effects ((point estimates and 95% confidence intervals) of one-point increases in the Policy stringency index (lagged with one week) on the log of the number of COVID-19 for the three waves of the pandemic in The Netherlands, 2020-2022. Results based on Model S2a in Table S2.

### Part IV: Excess mortality as a function of policy restrictions and mobility changes

Lastly, we look at excess mortality per week in each province as a function of the policy stringency index, changes in mobility and province fixed effects and demographic covariates. Policy stringency (lagged with 6 weeks) has significant negative associations with excess mortality. In terms of effect size, the one-step ahead direct effect of the imposition of a full societal lockdown (which is equivalent to going from 1 to 10 on the policy stringency index) is a reduction of excess mortality with approximately 2.2 percentage points (e.g. from the median of 4.7% to 2.5% excess mortality). In addition, we have the mediated effect of policy via mobility, which adds another sizable reduction (for example, looking at the coefficient for policy stringency on presence in work places from Model 2a in Table 2 [−2.74] and multiplying it by the effect of mobility in places of work from Model 4a in Table 4 [0.002] leads to an estimate of 0.5 percentage points reduction in excess mortality for a one-unit change in policy stringency).

**Table 4.**
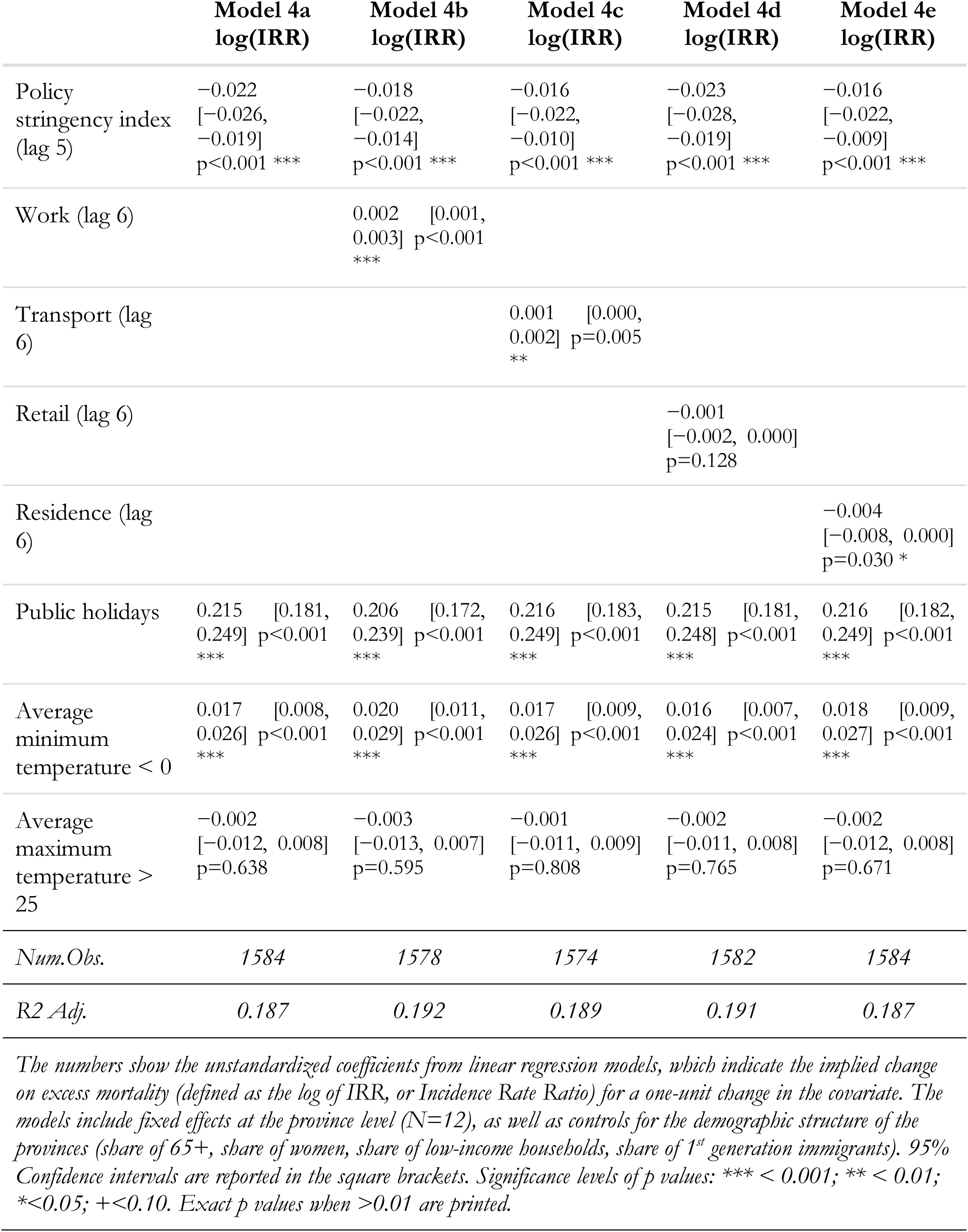
Linear regression models of excess mortality (the log of IRR, or the Incidence Rate Ratio).

|  | **Model 4a**<br>**log(IRR)** | **Model 4b**<br>**log(IRR)** | **Model 4c**<br>**log(IRR)** | **Model 4d**<br>**log(IRR)** | **Model 4e**<br>**log(IRR)** |
| --- | --- | --- | --- | --- | --- |
| Policy stringency index (lag 5) | -0.022<br>[-0.026, -0.019]<br>p<0.001 *** | -0.018<br>[-0.022, -0.014]<br>p<0.001 *** | -0.016<br>[-0.022, -0.010]<br>p<0.001 *** | -0.023<br>[-0.028, -0.019]<br>p<0.001 *** | -0.016<br>[-0.022, -0.009]<br>p<0.001 *** |
| Work (lag 6) |  | 0.002 [0.001, 0.003] p<0.001 *** |  |  |  |
| Transport (lag 6) |  |  | 0.001 [0.000, 0.002] p=0.005 ** |  |  |
| Retail (lag 6) |  |  |  | -0.001 [-0.002, 0.000] p=0.128 |  |
| Residence (lag 6) |  |  |  |  | -0.004 [-0.008, 0.000] p=0.030 * |
| Public holidays | 0.215 [0.181, 0.249] p<0.001 *** | 0.206 [0.172, 0.239] p<0.001 *** | 0.216 [0.183, 0.249] p<0.001 *** | 0.215 [0.181, 0.248] p<0.001 *** | 0.216 [0.182, 0.249] p<0.001 *** |
| Average minimum temperature < 0 | 0.017 [0.008, 0.026] p<0.001 *** | 0.020 [0.011, 0.029] p<0.001 *** | 0.017 [0.009, 0.026] p<0.001 *** | 0.016 [0.007, 0.024] p<0.001 *** | 0.018 [0.009, 0.027] p<0.001 *** |
| Average maximum temperature > 25 | -0.002 [-0.012, 0.008] p=0.638 | -0.003 [-0.013, 0.007] p=0.595 | -0.001 [-0.011, 0.009] p=0.808 | -0.002 [-0.011, 0.008] p=0.765 | -0.002 [-0.012, 0.008] p=0.671 |
| *Num.Obs.* | *1584* | *1578* | *1574* | *1582* | *1584* |
| *R2 Adj.* | *0.187* | *0.192* | *0.189* | *0.191* | *0.187* |

*The numbers show the unstandardized coefficients from linear regression models, which indicate the implied change on excess mortality (defined as the log of IRR, or Incidence Rate Ratio) for a one-unit change in the covariate. The models include fixed effects at the province level (N=12), as well as controls for the demographic structure of the provinces (share of 65+, share of women, share of low-income households, share of 1<sup>st</sup> generation immigrants). 95% Confidence intervals are reported in the square brackets. Significance levels of p values: \*\*\* < 0.001; \*\* < 0.01; \* < 0.05; + < 0.10. Exact p values when > 0.01 are printed.*

The evidence for separate effects of changes in mobility is mixed: presence in places of work and in transport hubs is associated with higher excess mortality according to Table 4, but the exact lag selected for the mobility variables matters substantially (see Figure 5 below). Expectedly, increased presence in places of residence is associated with lower excess mortality. But there is no evidence for a positive effect of presence in grocery shops and markets (retail).

From the control variables, it is noteworthy that colder weeks (with average minimum temperature lower than 0 degrees Celsius) have a significant positive association with excess mortality, while very warm weeks (with average maximum temperature above 25 degrees Celsius) do not. Public holidays are associated with higher excess mortality, even though they were not associated with more COVID-19 cases.

When we look at possible interaction effects, the policy effect on excess mortality is most pronounced during the first wave and gets smaller during the second and third waves (for details see Table S3 in the Supporting Information). The effect was greatest in the Southern provinces Limburg and North Brabant (see Table S3 and Figure 4) and it was smallest in the less densely populated provinces Drenthe, Flevoland and Zeeland.

**Fig 4.**
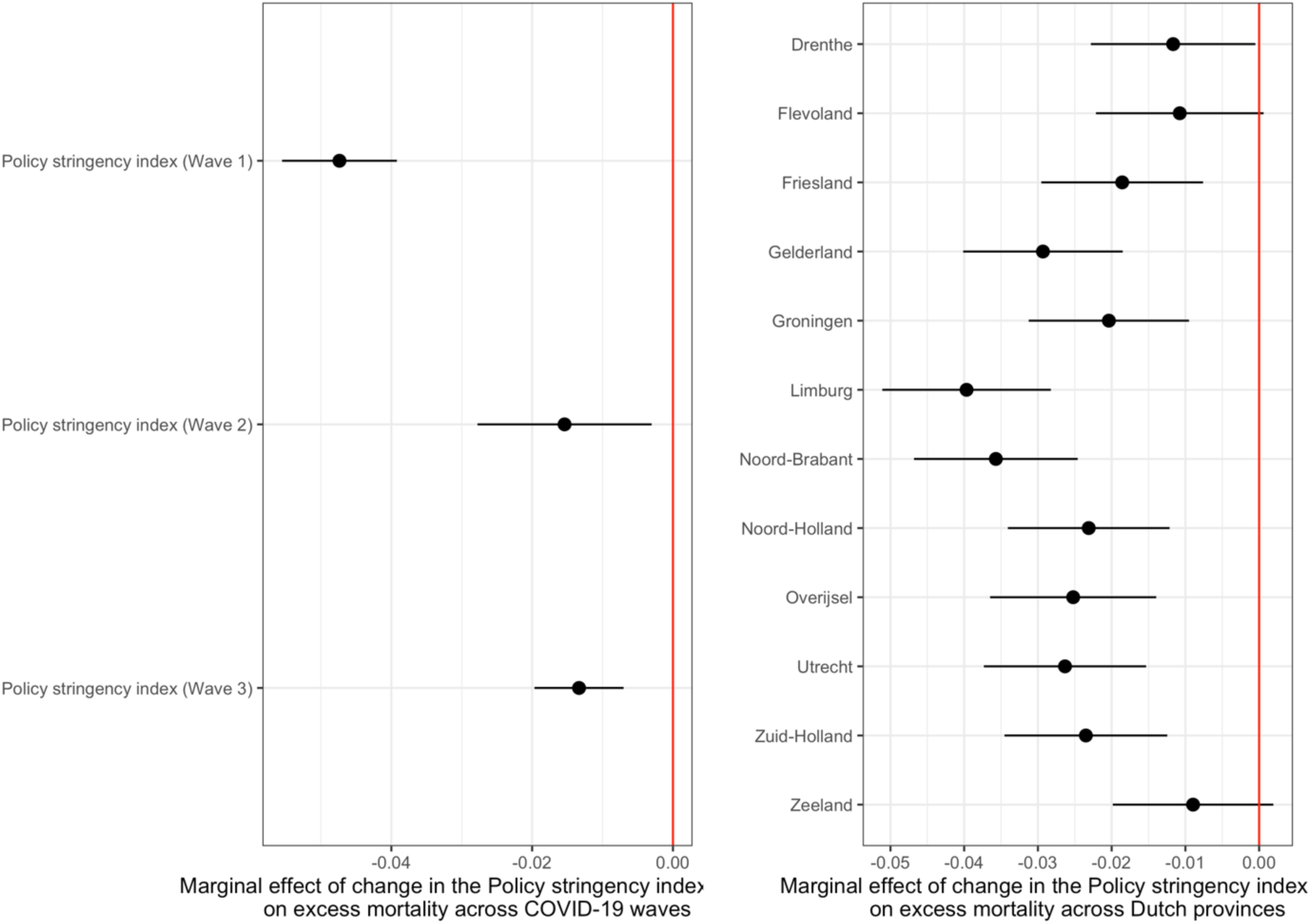
Marginal effects of changes in the Policy stringency index (lag 5) on excess mortality (log IRR) across COVID-19 waves (left panel) and across Dutch provinces (right panel), 2020-2022 The figure shows the predicted marginal effects ((point estimates and 95% confidence intervals) of one-point increases in the Policy stringency index (lagged with five weeks) on the log of the IRR (Incidence Rate Ratio) for the three waves of the pandemic (left panel) and for the 12 provinces (right panel) in The Netherlands, 2020-2022. Results based on the models reported in Tables S3 in the Supporting Information.

**Fig 5.**
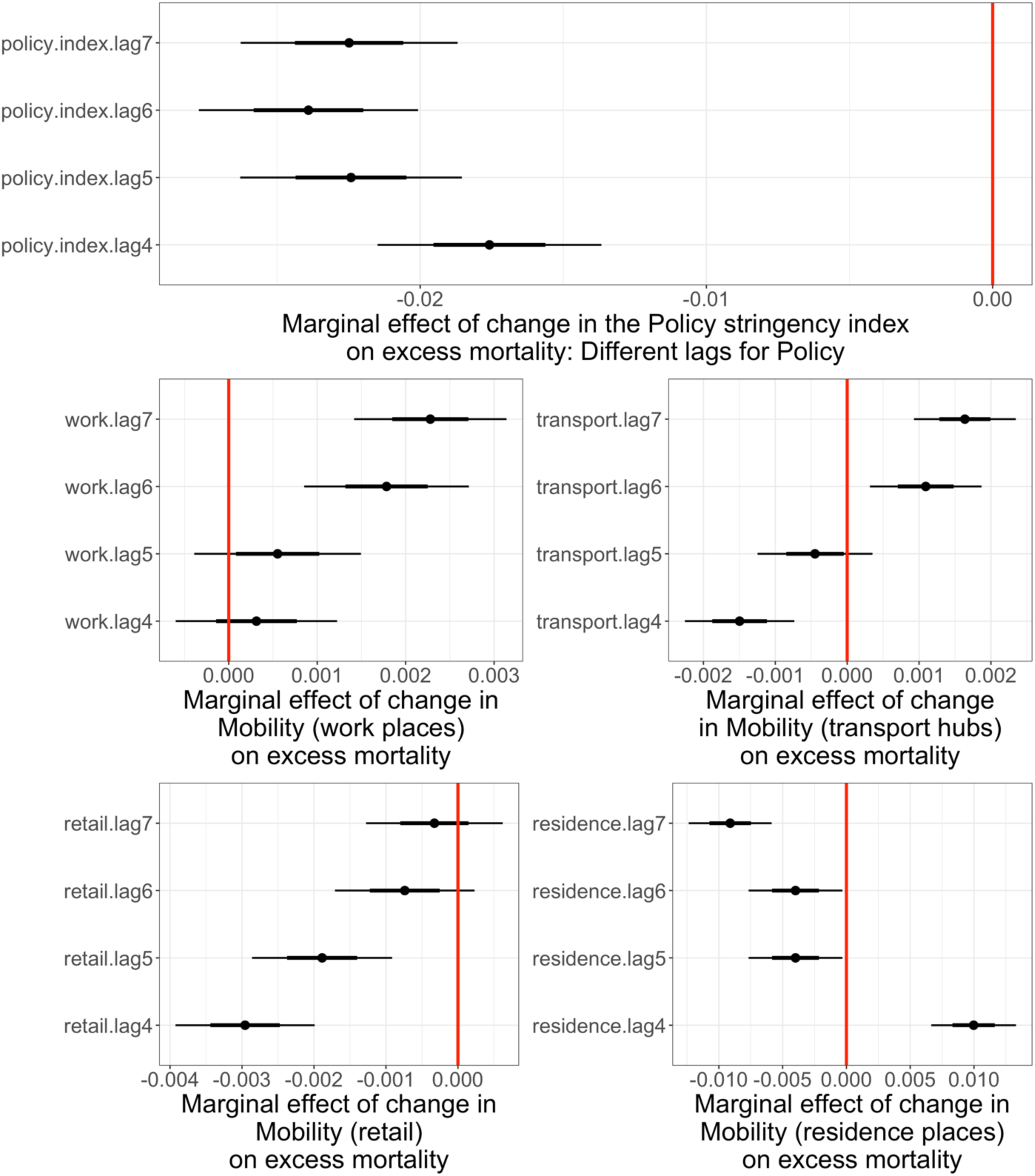
Marginal effects of Policy stringency and Mobility on excess mortality (log IRR) for different lags of the predictors, in The Netherlands, 2020-2022 The figure shows the predicted marginal effects ((point estimates and 95% confidence intervals) of one-point increases in the Policy stringency index (top panel) and different aspects of Mobility (bottom four panels) on excess mortality (defined as the log of the IRR, Incidence Rate Ratio) in The Netherlands, 2020-2022, for different lags of the predictors.

The results remain similar when we model the number of registered COVID-19 deaths instead of our estimates of excess mortality (see Table S4 in the Supporting Information): policy stringency continues to have a significant negative association with the number of COVID-19 deaths five weeks ahead, and the pattern of results for the effects of mobility is largely as the one in Table 4.

We also explore the sensitivity of these results to the exact specification of the lag structure of the predictors. To do so, we enter the policy restrictions and mobility variables with lags from 4 to 7 weeks. The results are shown in Figure 5. The coefficient of the policy variable is quite stable for different lags, especially for lags between 5 and 7. The coefficients for the changes in mobility are sensitive to the exact lag specification. In all models depicted in Figure 5, when the coefficient for Mobility varies, the coefficient for Policy is fixed to lag 5. When it comes to mobility related to workplaces, the expected positive coefficient is found in lags 6 and 7. For transport, the coefficient is also positive in lags 6 and 7, but negative in lag 4. For retail, with higher lags (6 and 7) the coefficient for mobility moves from negative to non-distinguishable from zero. For residence, the coefficient is negative and significant for lags 5 to 7.

Altogether, higher lags (6 and 7) deliver more significant results in the excepted direction. Again, we should emphasize that this is contingent on the policy being entered in the model with lag 5. Given the complex correlation over time between Policy and Mobility, other patterns of results for mobility might be observed with different lag of policy being accounted for in the model. The results are robust to including fixed effects for the calendar months. In fact, many of the coefficients of interest are estimated more precisely and the policy effects appear slightly bigger in size.

## Discussion and Conclusion

The study examined the impact of COVID-19 policy restrictions on excess mortality in The Netherlands for the 2020-2022 period. We theorized the mechanisms connecting COVID-19 cases and deaths, policy stringency, changes in mobility patterns, and excess mortality and modeled the relationships among these using data disaggregated weekly and at the province level. The focus on excess mortality in place of COVID-19 cases and related deaths alone (17,21), its analysis beyond the first wave to include later stages of the pandemic, and the inclusion of within-country variation (13,14) together mark the importance of our contribution.

We show that the number of COVID-19 cases and deaths is associated with the strictness of policy restrictions, which is no surprise, as policy makers were actively monitoring and responding to the information about the course of the pandemic. It is nevertheless informative to see a confirmation of this in the data, and the differential impact of the cases and deaths over the course of the pandemic, in particular.

Our results confirm the findings of previous research that policy restrictions, by reducing mobility, helped to curb the spread of COVID-19 and limit excess mortality (10). We found that COVID-19 cases and deaths led to more restrictive policies and that greater policy stringency in turn led to reductions in presence at workplaces, transportation hubs, and retail locations, while increasing presence at home. These changes in mobility patterns in turn were associated with lower COVID-19 case counts two weeks later. Ultimately, more stringent policies, by limiting mobility in areas with a potential for high transmission, led to lower excess mortality with a lag of six weeks.

Importantly, we observed significant variation in these relationships across the different waves of the pandemic. The impact of case numbers on the stringency of restrictions declined with each wave, while that of deaths was constant in the first two waves but increased in the third. Policymakers may have become more responsive to the most severe health outcomes as the pandemic continued, as the virus variant during the third wave was milder than the previous variants (30). The effect of policy stringency on mobility was strongest during the first wave and declined in subsequent waves. This may reflect ‘pandemic fatigue’ and reduced compliance over time. Relatedly, the impact of policy on the number of cases was most pronounced in the second wave.

Interestingly, we did not find a significant relationship between changes in presence in grocery stores and retail shops on COVID-19 case counts. This contrasts with the strong effects observed for workplace and transportation mobility. It may be that other preventive behaviors, such as mask-wearing (which became officially recommended during the second wave in the fall of 2020) and distancing, were important for reducing transmission in these essential retail settings. Overall, our findings highlight the critical role that policy-induced reductions in mobility played in mitigating the health impacts of COVID-19. However, the declining effectiveness of these measures over time, and the existence of direct policy effects beyond mobility, underscore the need for policymakers to continually assess and adapt their strategies as pandemics evolve. At the same time, policymakers must carefully weigh the benefits of restrictive measures against the significant costs they impose on personal liberty.

Further research into the effectiveness of different policy mixes is essential to guide pandemic responses and prepare for future public health emergencies. For example, it would be fruitful to see replications of our analytic approach in other countries with more regional variation in policy restrictions. Communicating the findings of such research so that it can help decision makers strike the necessary balance between public health objectives and individual rights is equally important. Responses to the next pandemic can benefit from valuables lessons learned from the experience of COVID-19 with respect to the effectiveness of government interventions and the limits to their effects.

## Data Availability

The data will be made available upon publication on the Harvard Dataverse.

## Acknowledgements

We would like to thank the members of the research team part of the project on excess mortality in The Netherlands financed by the Dutch organization for knowledge and innovation in health, healthcare and well-being (ZomMW): No 104.302.522.100.07 and 104.302.522.200.03.

## Ethical approval

This study was approved by the Scientific Committee of the Department of Clinical Epidemiology of the Leiden University Medical Center (protocol A0199) with a waiver of participant consent, because it used exclusively pre-existing, de-identified data, which the Dutch Statistics Office (CBS) is allowed to process by law (Wet op het Centraal Bureau voor de Statistiek, i.e. Law for the CBS).

## Footnotes

5 The growth rate is calculated as the log in the number of new COVID-19 cases registered in week *x* minus the log of the number of new COVID-19 cases registered in week *x-1.* We add one case to all weeks to avoid the log function returning-Infinity in the very few weeks with zero cases.

## Supporting Information

**Table S1.**
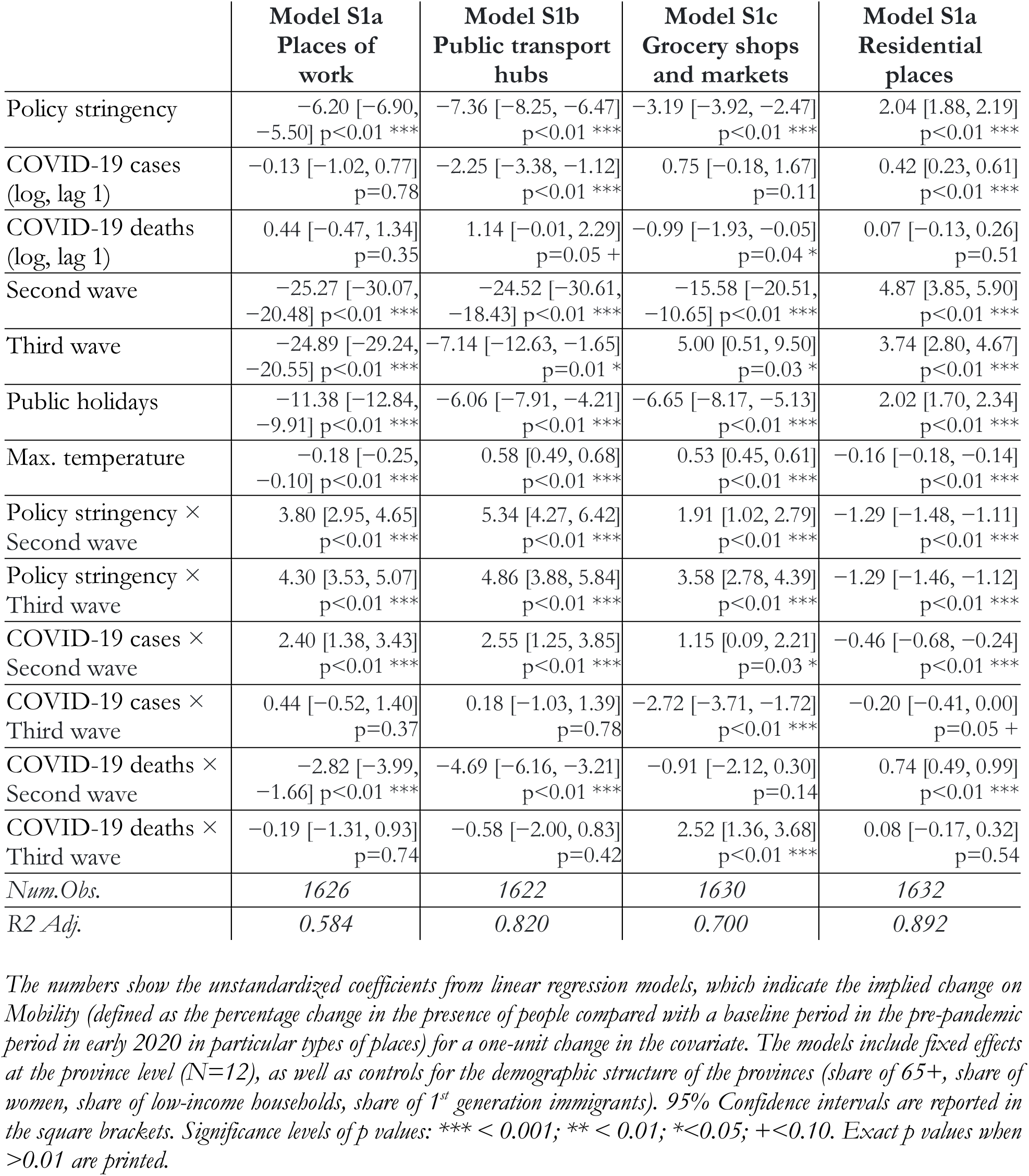
Changes in mobility as a function of policy stringency, COVID-19 cases and deaths, and additional covariates, including interaction effects with pandemic waves.

|  | **Model S1a**<br>**Places of**<br>**work** | **Model S1b**<br>**Public transport**<br>**hubs** | **Model S1c**<br>**Grocery shops**<br>**and markets** | **Model S1a**<br>**Residential**<br>**places** |
| --- | --- | --- | --- | --- |
| Policy stringency | -6.20 [-6.90, -5.50] p<0.01 *** | -7.36 [-8.25, -6.47] p<0.01 *** | -3.19 [-3.92, -2.47] p<0.01 *** | 2.04 [1.88, 2.19] p<0.01 *** |
| COVID-19 cases (log, lag 1) | -0.13 [-1.02, 0.77] p=0.78 | -2.25 [-3.38, -1.12] p<0.01 *** | 0.75 [-0.18, 1.67] p=0.11 | 0.42 [0.23, 0.61] p<0.01 *** |
| COVID-19 deaths (log, lag 1) | 0.44 [-0.47, 1.34] p=0.35 | 1.14 [-0.01, 2.29] p=0.05 + | -0.99 [-1.93, -0.05] p=0.04 * | 0.07 [-0.13, 0.26] p=0.51 |
| Second wave | -25.27 [-30.07, -20.48] p<0.01 *** | -24.52 [-30.61, -18.43] p<0.01 *** | -15.58 [-20.51, -10.65] p<0.01 *** | 4.87 [3.85, 5.90] p<0.01 *** |
| Third wave | -24.89 [-29.24, -20.55] p<0.01 *** | -7.14 [-12.63, -1.65] p=0.01 * | 5.00 [0.51, 9.50] p=0.03 * | 3.74 [2.80, 4.67] p<0.01 *** |
| Public holidays | -11.38 [-12.84, -9.91] p<0.01 *** | -6.06 [-7.91, -4.21] p<0.01 *** | -6.65 [-8.17, -5.13] p<0.01 *** | 2.02 [1.70, 2.34] p<0.01 *** |
| Max. temperature | -0.18 [-0.25, -0.10] p<0.01 *** | 0.58 [0.49, 0.68] p<0.01 *** | 0.53 [0.45, 0.61] p<0.01 *** | -0.16 [-0.18, -0.14] p<0.01 *** |
| Policy stringency × Second wave | 3.80 [2.95, 4.65] p<0.01 *** | 5.34 [4.27, 6.42] p<0.01 *** | 1.91 [1.02, 2.79] p<0.01 *** | -1.29 [-1.48, -1.11] p<0.01 *** |
| Policy stringency × Third wave | 4.30 [3.53, 5.07] p<0.01 *** | 4.86 [3.88, 5.84] p<0.01 *** | 3.58 [2.78, 4.39] p<0.01 *** | -1.29 [-1.46, -1.12] p<0.01 *** |
| COVID-19 cases × Second wave | 2.40 [1.38, 3.43] p<0.01 *** | 2.55 [1.25, 3.85] p<0.01 *** | 1.15 [0.09, 2.21] p=0.03 * | -0.46 [-0.68, -0.24] p<0.01 *** |
| COVID-19 cases × Third wave | 0.44 [-0.52, 1.40] p=0.37 | 0.18 [-1.03, 1.39] p=0.78 | -2.72 [-3.71, -1.72] p<0.01 *** | -0.20 [-0.41, 0.00] p=0.05 + |
| COVID-19 deaths × Second wave | -2.82 [-3.99, -1.66] p<0.01 *** | -4.69 [-6.16, -3.21] p<0.01 *** | -0.91 [-2.12, 0.30] p=0.14 | 0.74 [0.49, 0.99] p<0.01 *** |
| COVID-19 deaths × Third wave | -0.19 [-1.31, 0.93] p=0.74 | -0.58 [-2.00, 0.83] p=0.42 | 2.52 [1.36, 3.68] p<0.01 *** | 0.08 [-0.17, 0.32] p=0.54 |
| *Num.Obs.* | 1626 | 1622 | 1630 | 1632 |
| *R2 Adj.* | 0.584 | 0.820 | 0.700 | 0.892 |

*The numbers show the unstandardized coefficients from linear regression models, which indicate the implied change on Mobility (defined as the percentage change in the presence of people compared with a baseline period in the pre-pandemic period in early 2020 in particular types of places) for a one-unit change in the covariate. The models include fixed effects at the province level (N=12), as well as controls for the demographic structure of the provinces (share of 65+, share of women, share of low-income households, share of 1<sup>st</sup> generation immigrants). 95% Confidence intervals are reported in the square brackets. Significance levels of p values: \*\*\* < 0.001; \*\* < 0.01; \* < 0.05; + < 0.10. Exact p values when > 0.01 are printed.*

**Table S2.**
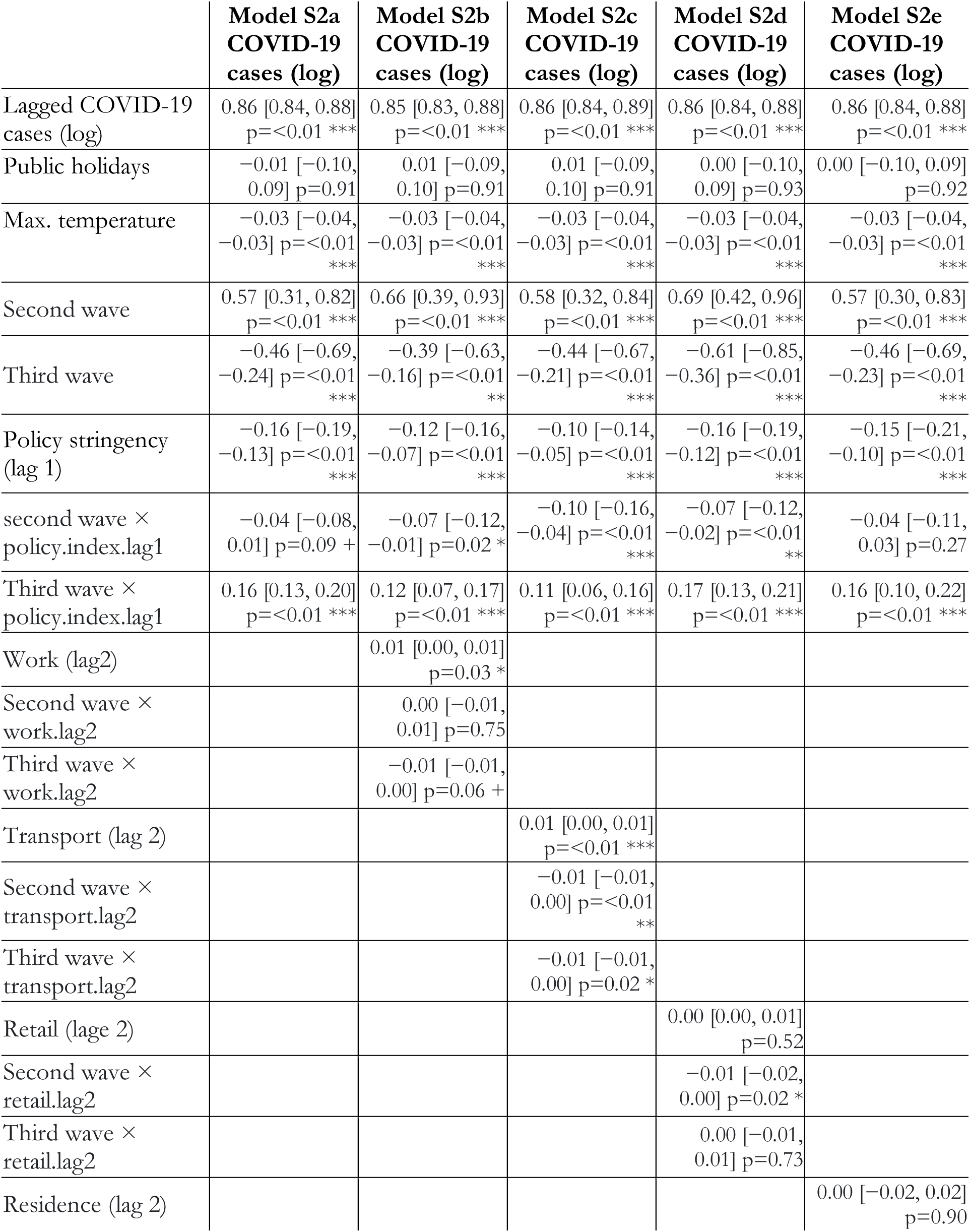
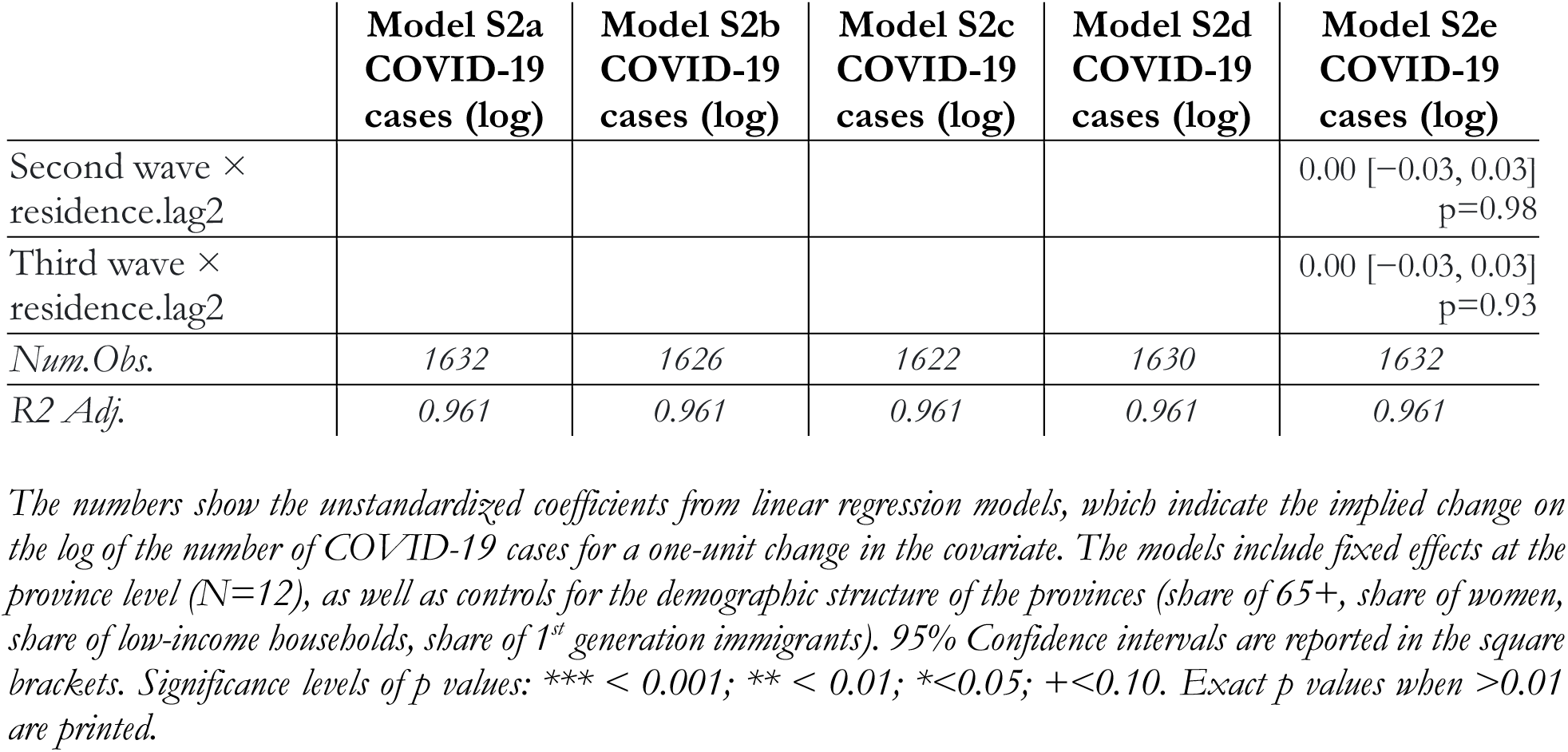
Number of registered COVID-19 cases (logged) as a function of policy stringency, changes in mobility and additional covariates, including interaction effects with pandemic waves.

**Table S3.** Linear regression models of excess mortality (the log of IRR, or the Incidence Rate Ratio), including interaction effects with pandemic waves and province.

|  | **Model S3a**<br>**log(IRR)** | **Model S3b**<br>**log(IRR)** |
| --- | --- | --- |
| Public holidays | 0.204 [0.170, 0.237]<br>p=<0.001 *** | 0.215 [0.181, 0.248]<br>p=<0.001 *** |
| Minimum temperature | 0.013 [0.005, 0.022]<br>p=0.003 ** | 0.017 [0.008, 0.025]<br>p=<0.001 *** |
| Maximum temperature | 0.000 [-0.010, 0.010]<br>p=0.997 | -0.003 [-0.013, 0.007]<br>p=0.535 |
| Policy stringency (lag 5) | -0.047 [-0.056, -0.039]<br>p=<0.001 *** | -0.012 [-0.023, -0.001]<br>p=0.041 * |
| Second wave | -0.146 [-0.225, -0.067]<br>p=<0.001 *** |  |
| Third wave | -0.144 [-0.205, -0.084]<br>p=<0.001 *** |  |
| Policy stringency (lag 5) × Second wave | 0.032 [0.017, 0.047]<br>p=<0.001 *** |  |
| Policy stringency (lag 5) × Third wave | 0.034 [0.024, 0.044]<br>p=<0.001 *** |  |
| Flevoland (FL) [vs. Drenthe (DR)] | 0.896 [0.006, 1.785]<br>p=0.048 * | 1.449 [0.626, 2.273]<br>p=<0.001 *** |
| Friesland (FR) | 1.703 [1.115, 2.291]<br>p=<0.001 *** | 2.496 [1.898, 3.095]<br>p=<0.001 *** |
| Gelderland (GE) | -0.195 [-0.439, 0.049] p=0.117 | -0.141 [-0.370, 0.088]<br>p=0.228 |
| Groningen (GR) | 2.881 [1.662, 4.099]<br>p=<0.001 *** | 4.118 [2.816, 5.420]<br>p=<0.001 *** |
| Limburg (LI) | 0.987 [0.475, 1.499]<br>p=<0.001 *** | 1.409 [0.821, 1.997]<br>p=<0.001 *** |
| Noord-Brabant (NB) | 1.074 [0.599, 1.550]<br>p=<0.001 *** | 1.684 [1.217, 2.152]<br>p=<0.001 *** |
| Noord-Holland (NH) | 0.343 [-0.549, 1.234]<br>p=0.451 | 0.527 [-0.380, 1.433]<br>p=0.255 |
| Overijssel (OV) | 1.420 [0.897, 1.943]<br>p=<0.001 *** | 2.182 [1.646, 2.719]<br>p=<0.001 *** |
| Utrecht (UT) | -1.353 [-2.024, -0.683]<br>p=<0.001 *** | -1.742 [-2.362, -1.123]<br>p=<0.001 *** |
| Zeeland (ZE) | 0.008 [-0.330, 0.346]<br>p=0.963 | -0.063 [-0.367, 0.241]<br>p=0.684 |
| Zuid-Holland (ZH) | 0.337 [-0.529, 1.202]<br>p=0.446 | 0.519 [-0.358, 1.396]<br>p=0.246 |
| FL × Policy stringency (lag 5) |  | 0.001 [-0.015, 0.016]<br>p=0.909 |
| FR × Policy stringency (lag 5) |  | -0.007 [-0.022, 0.008]<br>p=0.371 |
| GE × Policy stringency (lag 5) |  | -0.018 [-0.033, -0.002]<br>p=0.023 * |
| GR × Policy stringency (lag 5) |  | −0.009 [−0.024, 0.007]<br>p=0.264 |
| LI × Policy stringency (lag 5) |  | −0.028 [−0.044, −0.012]<br>p<0.001 *** |
| NB × Policy stringency (lag 5) |  | −0.024 [−0.040, −0.008]<br>p=0.002 ** |
| NH × Policy stringency (lag 5) |  | −0.011 [−0.027, 0.004]<br>p=0.151 |
| OV × Policy stringency (lag 5) |  | −0.014 [−0.029, 0.002]<br>p=0.080 + |
| UT × Policy stringency (lag 5) |  | −0.015 [−0.030, 0.001]<br>p=0.061 + |
| ZE × Policy stringency (lag 5) |  | 0.003 [−0.013, 0.018]<br>p=0.736 |
| ZH × Policy stringency (lag 5) |  | −0.012 [−0.027, 0.004]<br>p=0.135 |
| *Num.Obs.* | *1596* | *1596* |
| *R2 Adj.* | *0.196* | *0.184* |

The numbers show the unstandardized coefficients from linear regression models, which indicate the implied change on excess mortality (defined as the log of IRR, or Incidence Rate Ratio) for a one-unit change in the covariate. The models include fixed effects at the province level (N=12), as well as controls for the demographic structure of the provinces (share of 65+, share of women, share of low-income households, share of 1<sup>st</sup> generation immigrants). 95% Confidence intervals are reported in the square brackets. Significance levels of p values: \*\*\* < 0.001; \*\* < 0.01; \* < 0.05; + < 0.10. Exact p values when > 0.01 are printed.

**Table S4.** Number of registered COVID-19 deaths as a function of policy stringency, changes in mobility and additional covariates.

|  | Model S4a<br>N Deaths | Model S4b<br>N Deaths | Model S4c<br>N Deaths | Model S4d<br>N Deaths | Model S4e<br>N Deaths |
| --- | --- | --- | --- | --- | --- |
| Number of COVID-19 deaths (lag 1) | 0.820 [0.795, 0.844]<br>p=<0.001 *** | 0.818 [0.795, 0.842]<br>p=<0.001 *** | 0.812 [0.788, 0.836]<br>p=<0.001 *** | 0.822 [0.798, 0.846]<br>p=<0.001 *** | 0.818 [0.795, 0.842]<br>p=<0.001 *** |
| Public holidays | 0.980 [-1.505, 3.465] p=0.439 | 0.639 [-1.743, 3.022] p=0.599 | 1.125 [-1.219, 3.469] p=0.347 | 1.365 [-1.000, 3.731] p=0.258 | 0.929 [-1.407, 3.265] p=0.436 |
| Minimum temperature | -0.589 [-0.716, -0.463]<br>p=<0.001 *** | -0.542 [-0.665, -0.418]<br>p=<0.001 *** | -0.622 [-0.749, -0.496]<br>p=<0.001 *** | -0.567 [-0.702, -0.433]<br>p=<0.001 *** | -0.647 [-0.774, -0.519]<br>p=<0.001 *** |
| Second wave | 0.366 [-1.570, 2.303] p=0.711 | 0.609 [-1.295, 2.513] p=0.530 | 0.553 [-1.331, 2.436] p=0.565 | 1.002 [-0.905, 2.909] p=0.303 | -1.492 [-3.513, 0.530] p=0.148 |
| Third wave | -4.696 [-7.447, -1.946]<br>p=<0.001 *** | -3.467 [-6.178, -0.756]<br>p=0.012 * | -4.428 [-7.158, -1.699]<br>p=0.001 ** | -3.741 [-6.640, -0.842]<br>p=0.011 * | -6.345 [-9.195, -3.495]<br>p=<0.001 *** |
| Policy stringency index (lag 5) | -2.014 [-2.349, -1.679]<br>p=<0.001 *** | -1.428 [-1.791, -1.064]<br>p=<0.001 *** | -0.949 [-1.365, -0.534]<br>p=<0.001 *** | -1.682 [-2.022, -1.342]<br>p=<0.001 *** | -0.499 [-0.989, -0.009]<br>p=0.046 * |
| Work (lag 6) |  | 0.135 [0.072, 0.198]<br>p=<0.001 *** |  |  |  |
| Transport (lag 6) |  |  | 0.171 [0.116, 0.225]<br>p=<0.001 *** |  |  |
| Retail (lag 6) |  |  |  | 0.060 [-0.015, 0.135]<br>p=0.119 |  |
| Residence (lag 6) |  |  |  |  | -0.928 [-1.201, -0.654]<br>p=<0.001 *** |
| Num.Obs. | 1596 | 1578 | 1574 | 1582 | 1584 |
| R2 Adj. | 0.846 | 0.859 | 0.861 | 0.857 | 0.861 |

The numbers show the unstandardized coefficients from linear regression models, which indicate the implied change on the number of COVID-19 deaths for a one-unit change in the covariate. The models include fixed effects at the province level (N=12), as well as controls for the demographic structure of the provinces (share of 65+, share of women, share of low-income households, share of 1<sup>st</sup> generation immigrants). 95% Confidence intervals are reported in the square brackets. Significance levels of p values: \*\*\* < 0.001; \*\* < 0.01; \* < 0.05; + < 0.10. Exact p values when > 0.01 are printed.

**Fig. S1.**
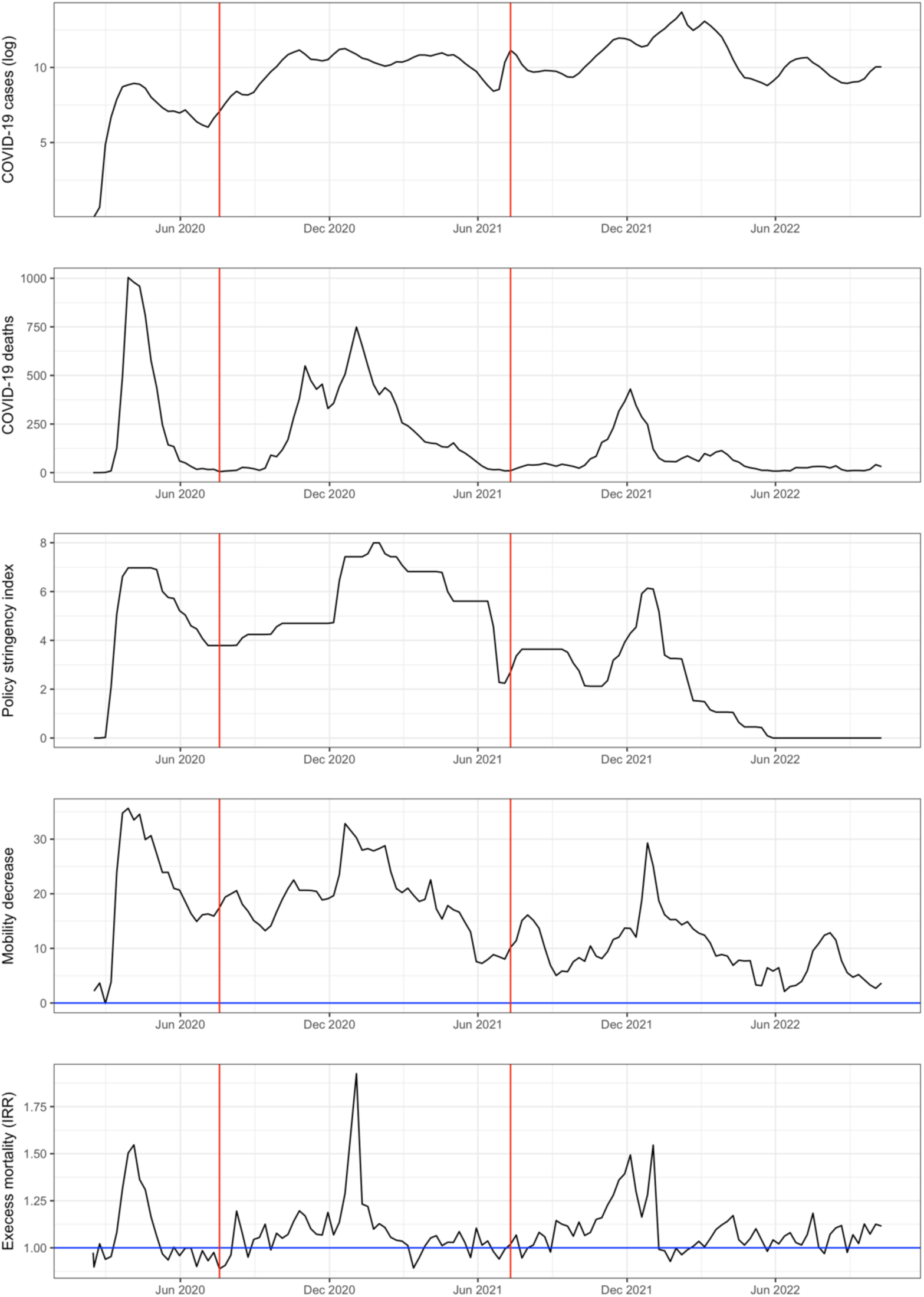
Descriptive trends of the main variables of interest Note: Weekly data between February 2020 and October 2022 in The Netherlands. The vertical red lines show when the second and third COVID-19 waves started. From top to bottom: log of registered COVID-19 cases; number of COVID-19 deaths; Policy stringency index (scaled between 0 and 10); Mobility decrease (average of four indexes from the Google Mobility Report for The Netherlands as whole; the original values have been scaled so that higher values indicate less presence in public places and more presence at home); Excess mortality (log of the IRR, or Incidence Rate Ratio, for The Netherlands as a whole).

**Fig. S2.**
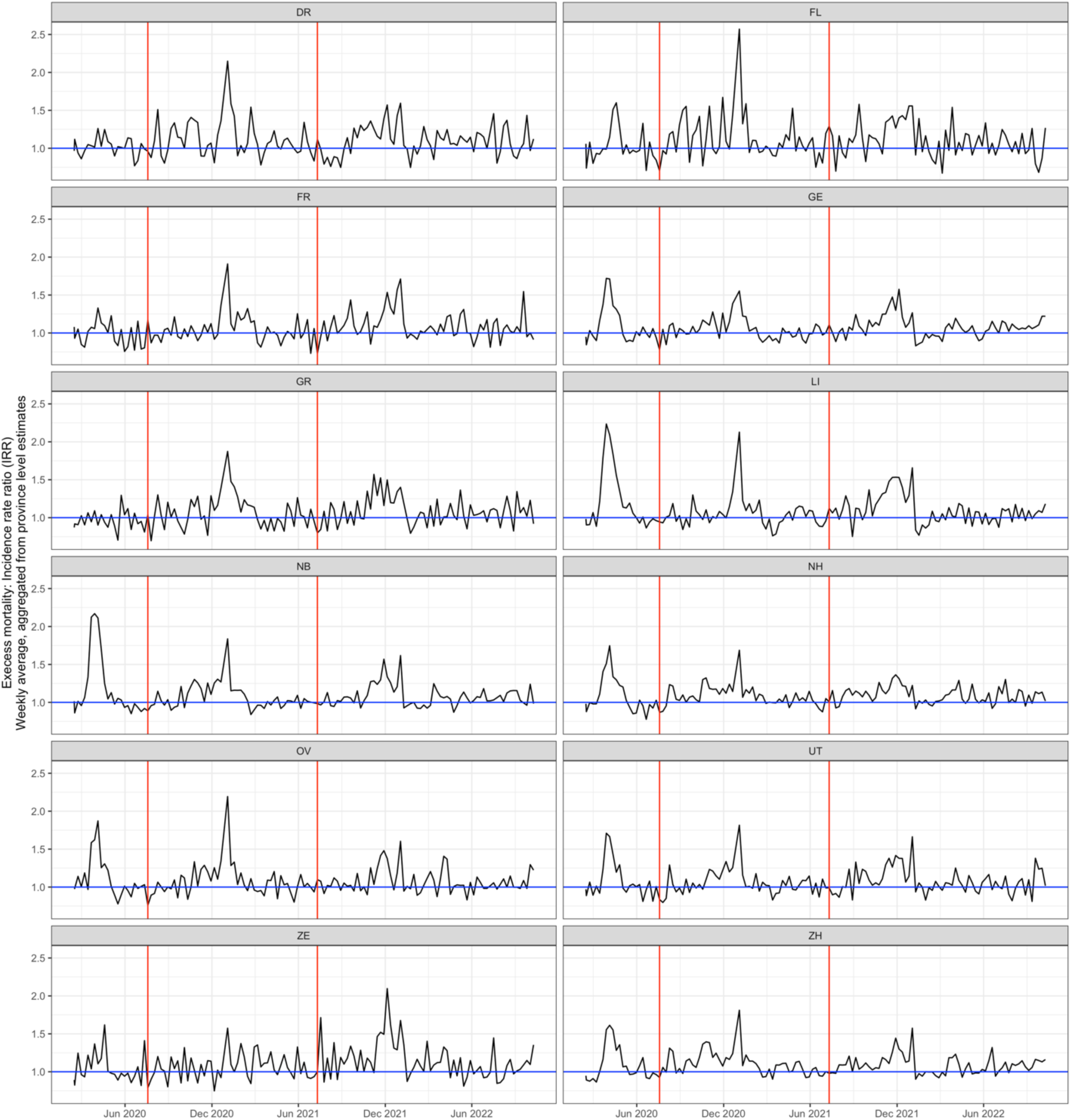
Excess mortality per province Note: Estimates of excess mortality (log IRR) in each of the twelve Dutch provinces. Weekly data between February 2020 and October 2022 in The Netherlands. The vertical red lines show when the second and third COVID-19 waves started. For province codes, see Table S3.

---

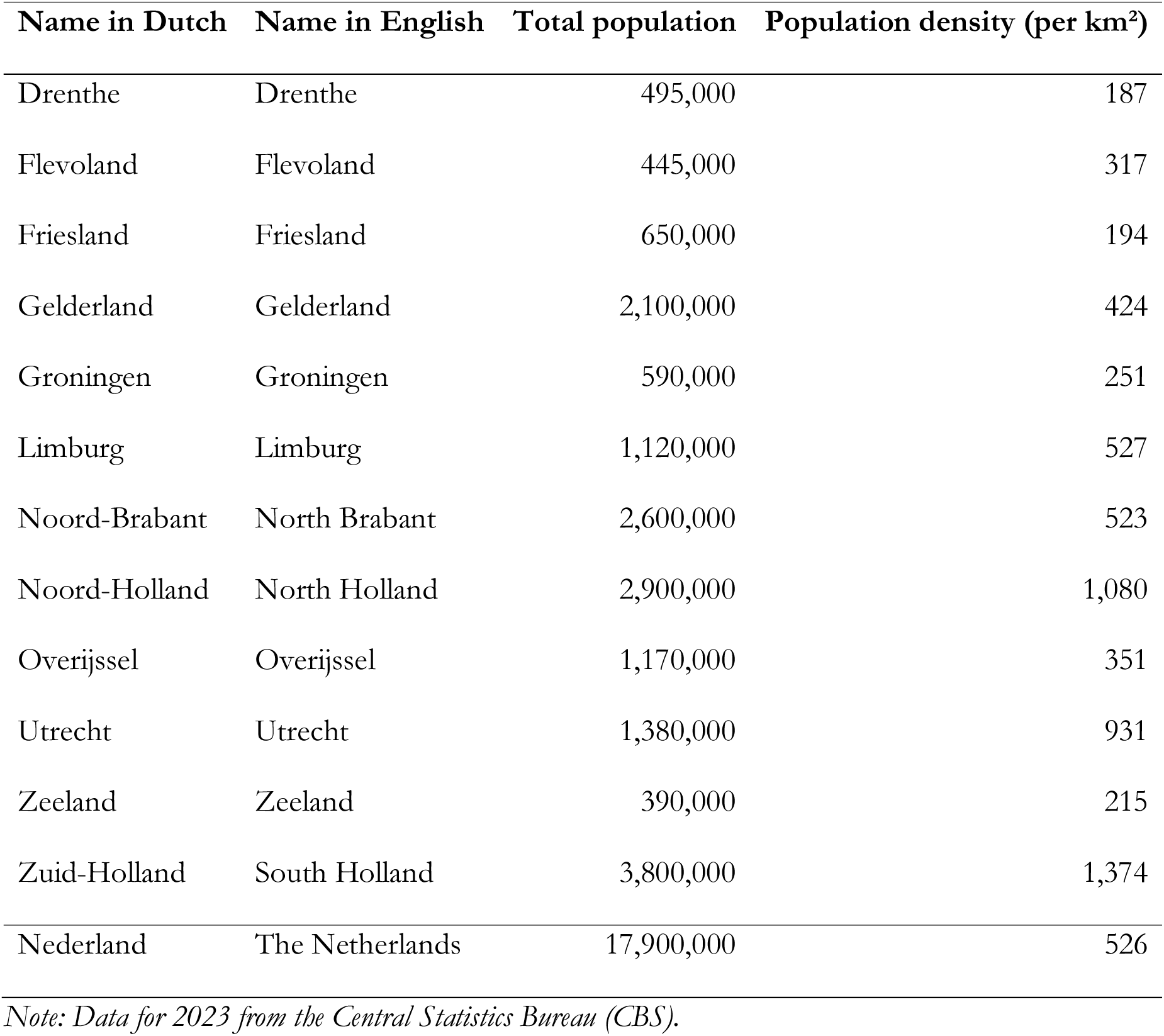
Information about the provinces in The Netherlands.

| **Name in Dutch** | **Name in English** | **Total population** | **Population density (per km<sup>2</sup>)** |
| --- | --- | --- | --- |
| Drenthe | Drenthe | 495,000 | 187 |
| Flevoland | Flevoland | 445,000 | 317 |
| Friesland | Friesland | 650,000 | 194 |
| Gelderland | Gelderland | 2,100,000 | 424 |
| Groningen | Groningen | 590,000 | 251 |
| Limburg | Limburg | 1,120,000 | 527 |
| Noord-Brabant | North Brabant | 2,600,000 | 523 |
| Noord-Holland | North Holland | 2,900,000 | 1,080 |
| Overijssel | Overijssel | 1,170,000 | 351 |
| Utrecht | Utrecht | 1,380,000 | 931 |
| Zeeland | Zeeland | 390,000 | 215 |
| Zuid-Holland | South Holland | 3,800,000 | 1,374 |
| Nederland | The Netherlands | 17,900,000 | 526 |

*Note: Data for 2023 from the Central Statistics Bureau (CBS).*

Acknowledgements

This article is a result of a broader research project in which the following team participated: Research group team (alphabetical order per institution):

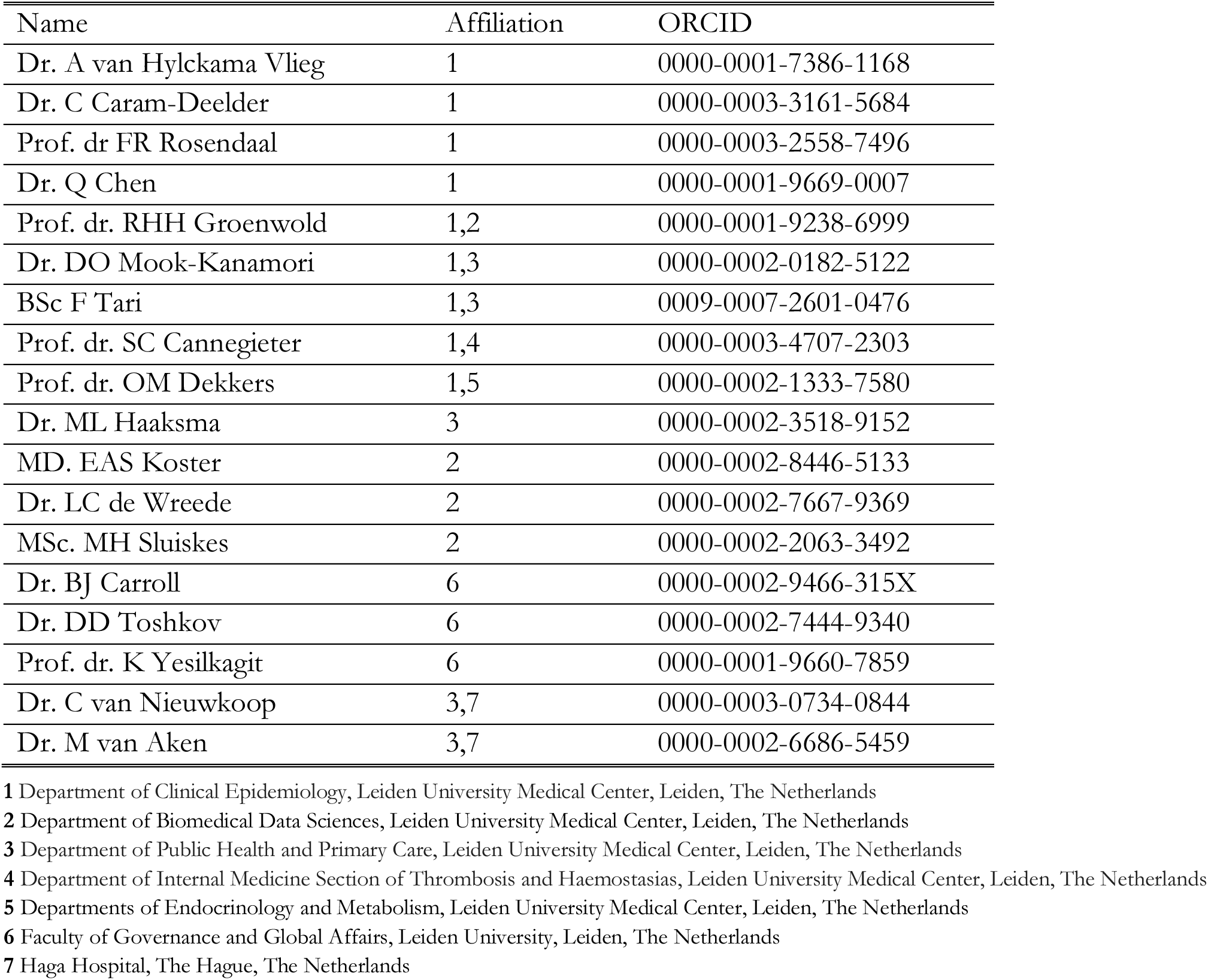

## Notes

### Competing Interest Statement

The authors have declared no competing interest.

## References

1. Hale T, Angrist N, Goldszmidt R, Kira B, Petherick A, Phillips T, et al. A global panel database of pandemic policies (Oxford COVID-19 Government Response Tracker). Nature human behaviour. 2021;5(4):529–38.

2. Chernozhukov V, Kasahara H, Schrimpf P. Causal impact of masks, policies, behavior on early covid-19 pandemic in the U.S. Journal of Econometrics. 2021;220(1):23–62.

3. Chernozhukov V, Kasahara H, Schrimpf P. The association of opening K–12 schools with the spread of COVID-19 in the United States: County-level panel data analysis. Proceedings of the National Academy of Sciences. 2021;118(42):e2103420118.

4. Méndez-Lizárraga CA, Castañeda-Cediel Ml, Delgado-Sánchez G, Ferreira-Guerrero EE, Ferreyra-Reyes L, Canizales-Quintero S, et al. Evaluating the impact of mobility in COVID-19 incidence and mortality: A case study from four states of Mexico. Frontiers in public health. 2022;10:877800.

5. Askitas N, Tatsiramos K, Verheyden B. Estimating worldwide effects of non-pharmaceutical interventions on COVID-19 incidence and population mobility patterns using a multiple-event study. Scientific Reports. 2021;11(1):1972.

6. Pozo-Martin F, Weishaar H, Cristea F, Hanefeld J, Bahr T, Schaade L, et al. The impact of non-pharmaceutical interventions on COVID-19 epidemic growth in the 37 OECD member states. Eur J Epidemiol. 2021 Jun 1;36(6):629–40.

7. Auger KA, Shah SS, Richardson T, Hartley D, Hall M, Warniment A, et al. Association Between Statewide School Closure and COVID-19 Incidence and Mortality in the US. JAMA. 2020 Sep 1;324(9):859–70.

8. Agrawal V, Cantor J, Sood N, Whaley C. The impact of COVID-19 shelter-in-place policy responses on excess mortality. Health Economics. 2023;32(11):2499–515.

9. White ER, Hébert-Dufresne L. State-level variation of initial COVID-19 dynamics in the United States. PLOS ONE. 2020 Oct 13;15(10):e0240648.

10. Hale T, Angrist N, Hale AJ, Kira B, Majumdar S, Petherick A, et al. Government responses and COVID-19 deaths: Global evidence across multiple pandemic waves. PLOS ONE. 2021 Jul 9;16(7):e0253116.

11. Haug N, Geyrhofer L, Londei A, Dervic E, Desvars-Larrive A, Loreto V, et al. Ranking the effectiveness of worldwide COVID-19 government interventions. Nat Hum Behav. 2020 Dec;4(12):1303–12.

12. Cho SW (Stanley). Quantifying the impact of nonpharmaceutical interventions during the COVID-19 outbreak: The case of Sweden. The Econometrics Journal. 2020 Sep 1;23(3):323–44.

13. Mendez-Brito A, El Bcheraoui C, Pozo-Martin F. Systematic review of empirical studies comparing the effectiveness of non-pharmaceutical interventions against COVID-19. Journal of Infection. 2021;83(3):281–93.

14. Perra N. Non-pharmaceutical interventions during the COVID-19 pandemic: A review. Physics Reports. 2021 May 23;913:1–52.

15. Herby J, Jonung L, Hanke SH. Were COVID-19 lockdowns worth it? A meta-analysis. Public Choice [Internet]. 2024 Nov 28 [cited 2025 Mar 12]; Available from: 10.1007/s11127-024-01216-7

16. Bonnet F, Grigoriev P, Sauerberg M, Alliger I, Mühlichen M, Camarda CG. Spatial disparities in the mortality burden of the covid-19 pandemic across 569 European regions (2020-2021). Nat Commun. 2024 May 18;15(1):4246.

17. Beaney T, Clarke JM, Jain V, Golestaneh AK, Lyons G, Salman D, et al. Excess mortality: the gold standard in measuring the impact of COVID-19 worldwide? J R Soc Med. 2020 Sep 1;113(9):329–34.

18. Karlinsky A, Kobak D. Tracking excess mortality across countries during the COVID-19 pandemic with the World Mortality Dataset. Davenport MP, Lipsitch M, Lipsitch M, Simonsen L, Mahmud A, editors. eLife. 2021 Jun 30;10:e69336.

19. Aburto JM, Schöley J, Kashnitsky I, Zhang L, Rahal C, Missov TI, et al. Quantifying impacts of the COVID-19 pandemic through life-expectancy losses: a population-level study of 29 countries. International Journal of Epidemiology. 2022 Feb 1;51(1):63–74.

20. Dorn F, Lange B, Braml M, Gstrein D, Nyirenda JLZ, Vanella P, et al. The challenge of estimating the direct and indirect effects of COVID-19 interventions – Toward an integrated economic and epidemiological approach. Economics & Human Biology. 2023 Apr 1;49:101198.

21. Caram-Deelder C, Vlieg A van H, Groenwold RHH, Chen Q, Mook-Kanamori DO, Dekkers OM, et al. Excess mortality during the first 2 years of the COVID-19 pandemic (2020-2021) in the Netherlands: Overall and across demographic subgroups. IJID Regions. 2025 Mar 1;14:100500.

22. Lison A, Banholzer N, Sharma M, Mindermann S, Unwin HJT, Mishra S, et al. Effectiveness assessment of non-pharmaceutical interventions: lessons learned from the COVID-19 pandemic. The Lancet Public Health. 2023 Apr 1;8(4):e311–7.

23. Badr HS, Du H, Marshall M, Dong E, Squire MM, Gardner LM. Association between mobility patterns and COVID-19 transmission in the USA: a mathematical modelling study. The Lancet Infectious Diseases. 2020;20(11):1247–54.

24. Schöley J, Aburto JM, Kashnitsky I, Kniffka MS, Zhang L, Jaadla H, et al. Life expectancy changes since COVID-19. Nat Hum Behav. 2022 Dec;6(12):1649–59.

25. Constantinou A, Kitson NK, Liu Y, Chobtham K, Amirkhizi AH, Nanavati PA, et al. Open problems in causal structure learning: A case study of COVID-19 in the UK. Expert Systems with Applications. 2023 Dec 30;234:121069.

26. Hale T, Angrist N, Goldszmidt R, Kira B, Petherick A, Phillips T, et al. A global panel database of pandemic policies (Oxford COVID-19 Government Response Tracker). Nat Hum Behav. 2021 Apr;5(4):529–38.

27. Berge L, Krantz S, McDermott G, Lenth R. Package ‘fixest.’ 2021;

28. Arel-Bundock V. modelsummary: Data and Model Summaries in R. Journal of Statistical Software. 2022;103:1–23.

29. Lander JP. coefplot: Plots Coefficients from Fitted Models. R package version 1.2. 4. 2016;

30. Yuan Z, Shao Z, Ma L, Guo R. Clinical Severity of SARS-CoV-2 Variants during COVID-19 Vaccination: A Systematic Review and Meta-Analysis. Viruses. 2023 Sep 26;15(10):1994.

